# Epigenome-wide Association Study of Alcohol Use Disorder in Five Brain Regions

**DOI:** 10.1101/2021.08.01.21261118

**Authors:** Lea Zillich, Josef Frank, Fabian Streit, Marion M Friske, Jerome C Foo, Lea Sirignano, Stefanie Heilmann-Heimbach, Helene Dukal, Franziska Degenhardt, Per Hoffmann, Anita C Hansson, Markus M Nöthen, Marcella Rietschel, Rainer Spanagel, Stephanie H Witt

**Author notes:** Corresponding author: Rainer Spanagel, Central Institute of Mental Health, Institute for Psychopharmacology, J5, 68159 Mannheim, Germany. shared senior authorship (Rainer Spanagel, Stephanie H Witt).

## Abstract

Alcohol Use Disorder (AUD) is closely linked to the brain regions forming the neurocircuitry of addiction. Postmortem human brain tissue enables the direct study of the molecular pathomechanisms of AUD. This study aims to identify these mechanisms by examining differential DNA-methylation between cases with severe AUD (n=53) and controls (n=58) using a brain region-specific approach, in which sample sizes ranged between 46 and 94. Samples of the anterior cingulate cortex (ACC), Brodmann Area 9 (BA9), caudate nucleus (CN), ventral striatum (VS), and putamen (PUT) were investigated. DNA-methylation levels were determined using the Illumina HumanMethylationEPIC Beadchip. Epigenome-wide association analyses were carried out to identify differentially methylated CpG-sites and regions between cases and controls in each brain region. Weighted Correlation Network Analysis (WGCNA), gene-set and GWAS-enrichment analyses were performed. Two differentially methylated CpG-sites were associated with AUD in the CN, and 18 in VS (*q* <.05). No epigenome-wide significant CpG-sites were found in BA9, ACC, or PUT. Differentially methylated regions associated with AUD case-/control status (*q* < .05) were found in the CN (n=6), VS (n=18) and ACC (n=1). In the VS, the WGCNA-module showing the strongest association with AUD was enriched for immune-related pathways. This study is the first to analyze methylation differences between AUD cases and controls in multiple brain regions and consists of the largest sample to date. Several novel CpG-sites and regions implicated in AUD were identified, providing a first basis to explore epigenetic correlates of AUD.

## Introduction

Every year, approximately 5.3% of all deaths worldwide are a result of the harmful use of alcohol and approximately 230 diseases are associated with alcohol use [1]. The lifetime prevalence of alcohol use disorder (AUD) varies globally, with North African/Middle Eastern countries having the lowest (0.59%) and Eastern European countries the highest (4.25%) prevalence. With a global prevalence of 1.32%, AUD is an important contributor to global disease burden [2]. AUD is a moderately heritable disease; a meta-analysis of twin studies estimated a heritability of 49% [3].

It has been proposed that drug-induced alterations in gene expression in the neurocircuitry of the brain contribute to addiction [4]. Recent evidence suggests that alterations in DNA-methylation, an epigenetic mechanism affecting gene expression, play an important role in addiction (for reviews see: [5,6]). Differential DNA-methylation is associated with alcohol consumption and AUD both in peripheral blood and postmortem brain tissue (for an overview see: Wedemeyer, et al. ^7^). Examining alterations in DNA-methylation in epigenome-wide association studies (EWAS) allows for the investigation of inter-individual differences which are attributable to a phenotype [8]. For example, a recent EWAS of AUD in peripheral blood suggests that networks in glucocorticoid signaling and inflammation-related genes are associated with AUD [9].

Human postmortem brain tissue is a sparse and valuable resource and allows a more direct characterization of AUD mechanisms than possible by analyzing peripheral blood [10]. So far, a small number of postmortem brain studies have been conducted, mostly investigating the prefrontal cortex (PFC), which, due to its role in reward regulation and higher-order executive function, is thought to be disrupted in addiction [11]. An EWAS comparing individuals with AUD with age-matched controls detected a range of differentially methylated CpG-sites in Brodmann Area 9 (BA9) in 16 pairs of males, but not in seven pairs of females [12]. Another study identified AUD-associated differentially methylated CpG-sites in Brodmann Area 10, which did not remain significant after multiple testing correction [13]. However, downstream analyses implicated *NR3C1*, a gene coding for the glucocorticoid receptor, which is crucial to stress regulation and found to be functionally relevant in AUD. The increased DNA-methylation in individuals with AUD was also associated with reduced *NR3C1* mRNA and protein expression levels [13].

Investigating DNA-methylation in the wider addiction neurocircuitry may give deeper insights into the pathophysiological mechanisms of AUD, and may reveal potential targets for treatment or prevention [14,15]. Dysfunction in the addiction neurocircuitry, which comprises areas involved in cognitive control such as the dorsolateral PFC, the anterior cingulate cortex (ACC), and regions in the basal ganglia, can have impairing consequences associated with disrupted reward-related decision-making, alcohol craving, and compulsive alcohol consumption [11,16]. Of particular interest is the striatum, which is divided into ventral and dorsal subdivisions based on function and connectivity. The ventral striatum (VS), comprises the nucleus accumbens (NAcc) and olfactory tubercle while the dorsal striatum contains the caudate nucleus (CN) and putamen (PUT) [17]. The NAcc is thought to be important in addiction due to its role in processing motivation, more precisely aversion and reward [17]. The caudate nucleus and putamen both influence motor function; in addition, the caudate is involved in goal-directed action, executive functioning and cognitive control, while the putamen is implicated in various types of learning, including reinforcement learning and habit formation [18]. In a study investigating DNA-methylation in PFC and NAcc, CpG-sites in *DLGAP2* emerged as differentially methylated between 39 male AUD cases and 47 controls in both brain regions; the differences were genotype-dependent [19].

In the present study, we aimed to identify epigenetic mechanisms associated with AUD, in five brain regions previously implicated in the neurocircuitry of addiction [17]. Brain-region specific EWAS of AUD were performed in the BA9, ACC, VS, CN and PUT.

## Materials and Methods

### Samples

In total, 395 human postmortem brain samples from 111 subjects (53 AUD, 58 controls) were obtained from the New South Wales Tissue Resource Center (University of Sydney, Australia) under study reference number 2009-238N-MA by the Ethics Committee II of the Medical Faculty Mannheim. AUD and control subjects were matched by age and sex. All individuals met the following inclusion criteria, which were determined by next-of-kin interviews: age >18, no history of severe psychiatric, neurodevelopmental, or other substance use disorders (except nicotine use disorder), and Western European ancestry. Individuals with AUD were classified according to DSM-IV criteria and had consumed at least 80g alcohol daily, whereas controls had consumed less than 20g. Methylation data was generated in two batches and each batch was analyzed separately. The first batch comprised 220 samples of BA9, ACC, CN, and VS from 28 cases and 27 controls. In the second batch, 175 samples from 56 additional individuals from the CN, VS, and PUT were analyzed. Material from one to five brain regions was available for each individual. Therefore, the sample composition varies between the brain region-specific analyses. A sample description can be found in Table 1. Table 2 shows the number of samples for each brain region and each batch. Additional phenotype information, such as cause of death and detailed exclusion criteria can be found in the Supplementary Information (Table S1 and Text S1).

**Table 1.**
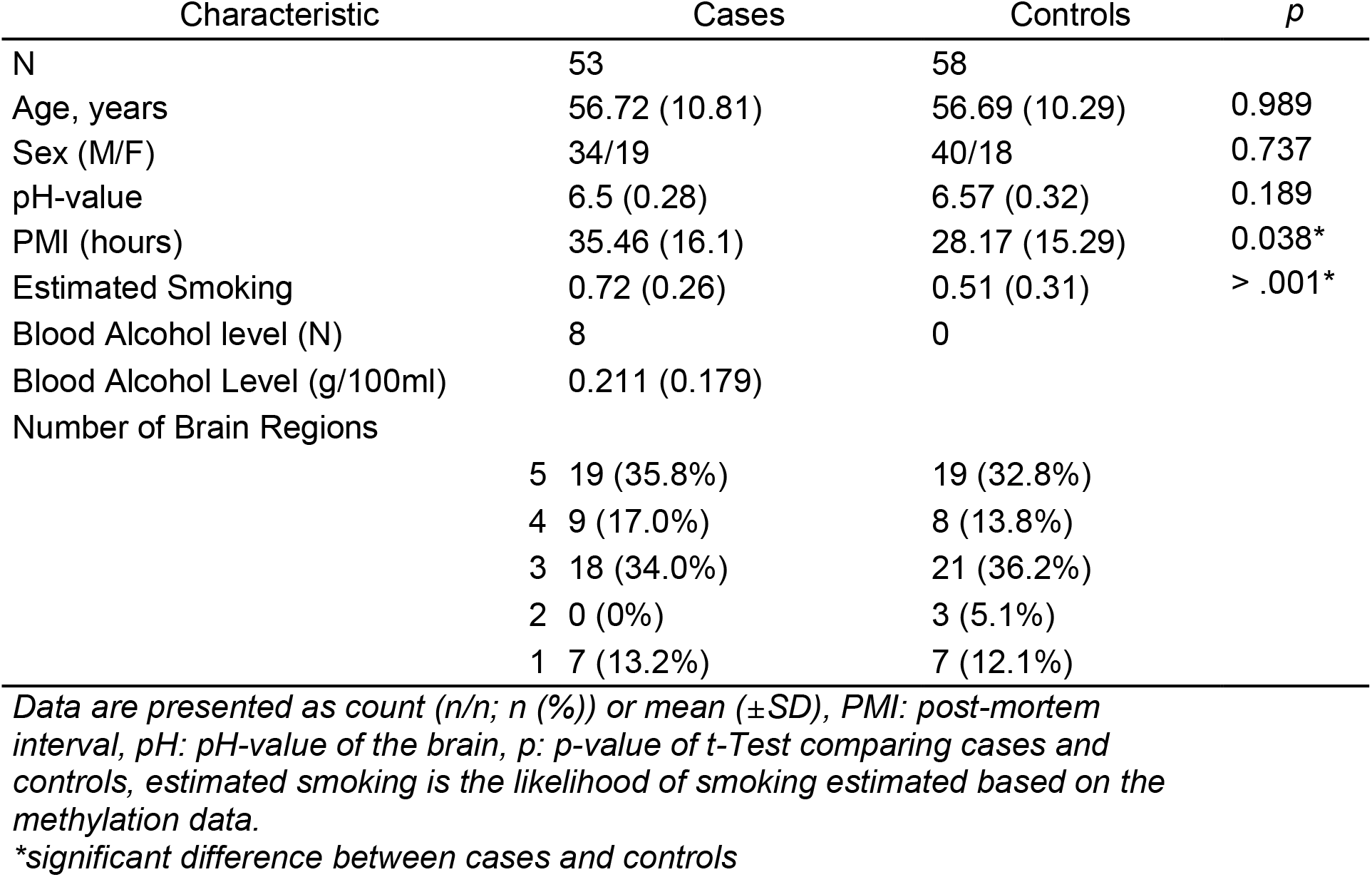
Descriptive statistics of demographic data.

**Table 2.**
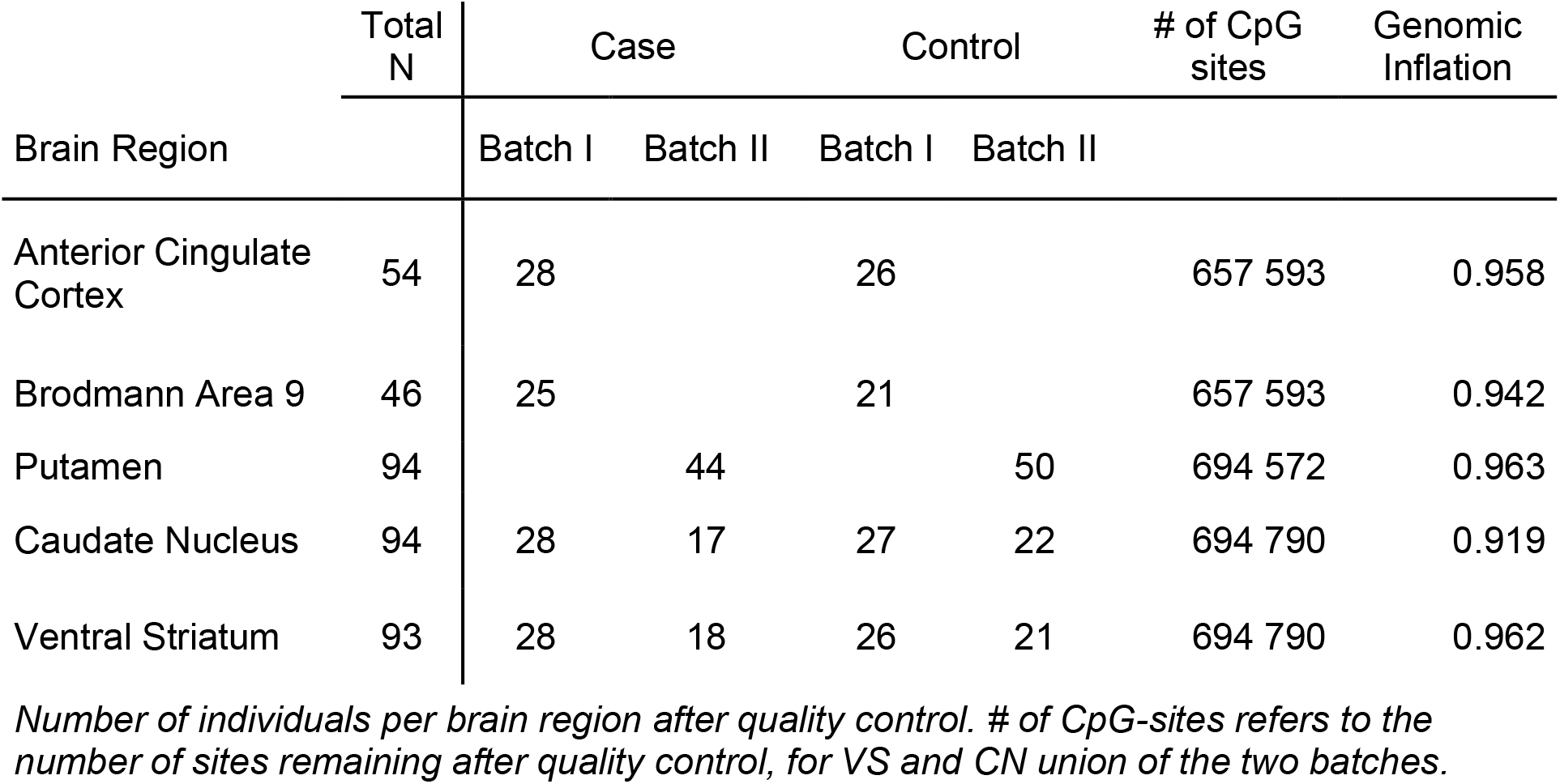
Sample Overview.

| Brain Region | Total N | Case |  | Control |  | # of CpG sites | Genomic Inflation |
| --- | --- | --- | --- | --- | --- | --- | --- |
|  |  | Batch I | Batch II | Batch I | Batch II |  |  |
| Anterior Cingulate Cortex | 54 | 28 |  | 26 |  | 657 593 | 0.958 |
| Brodmann Area 9 | 46 | 25 |  | 21 |  | 657 593 | 0.942 |
| Putamen | 94 |  | 44 |  | 50 | 694 572 | 0.963 |
| Caudate Nucleus | 94 | 28 | 17 | 27 | 22 | 694 790 | 0.919 |
| Ventral Striatum | 93 | 28 | 18 | 26 | 21 | 694 790 | 0.962 |

*Number of individuals per brain region after quality control. # of CpG-sites refers to the number of sites remaining after quality control, for VS and CN union of the two batches.*

### Epigenome-wide methylation

DNA was extracted from bulk brain tissue using the DNeasy extraction kit from Qiagen (Qiagen, Hilden, Germany). The genomic DNA samples were stored at -20°C. For the microarray analysis, the samples were randomized based on AUD case/control status and sex, and pipetted on processing plates. Due to the sample and different group sizes, samples from each brain region were processed on separate plates. Epigenome-wide methylation levels were determined using the Illumina HumanMethylationEPIC Beadchip and Illumina HiScan array scanning systems (Illumina, San Diego, CA).

### Data preprocessing, quality control, and filtering

All data preprocessing and analysis steps were performed using the R statistical environment, version 3.6.1. An updated version of the CPACOR-pipeline published by Lehne, et al. ^20^ was used to extract methylation data from raw intensity data and perform quality control. Samples were removed if (i) DNA quality was not sufficient (missing rate > 0.10) or (ii) a discrepancy between methylation-based and phenotypic sex emerged. Probes were removed when (i) the call-rate was insufficient (< 0.95), (ii) SNPs with a minor allele frequency > 0.10 were located in the probe sequence, (iii) the probes were located on the X or Y chromosome. After quality control 381 samples remained. Depending on the brain region, 657 593 – 694 791 sites were available for analysis after filtering. Detailed descriptions of sample size, the number of sites remaining after QC, and the inflation coefficient lambda for each model can be found in Table 2.

### Statistical Analysis

Methylation values were log-transformed (base 2) and included as dependent variables in the association analyses, as recommended by Du, et al. ^21^. Control for batch effects and technical quality was applied by extracting signals of the internal control probes of the EPIC array, performing principal component analysis (PCA), and extracting the first ten principal components. These were included as covariates in all association tests. To control for cell-type heterogeneity, cell counts were estimated using the method by Houseman, et al. ^22^, with the dorsolateral prefrontal cortex reference data [23]. This approach results in two estimates, one for neurons and one for other cell types. These were standardized so that the sum of both counts added up to one. The estimate for neurons was included as a covariate in all analyses.

Data on smoking was not available for all participants (missing for n=11, 10.81%). Smoking status was therefore estimated based on a validated set of sites [24]. Estimated smoking was included as a continuous covariate. 86% of current smokers were correctly classified; according to the regression model their likelihood of smoking was >50%.

#### Epigenome-wide association analysis

Tests of methylation differences between individuals with AUD and control subjects were performed with linear models, adjusting for sex, age, postmortem interval (PMI), pH-value, estimated smoking, standardized neuronal cell count, and the first ten principal components of the internal control probes. Each region and each batch was analyzed separately. The summary statistics for CN and VS were then meta-analyzed based on effect estimates and standard errors using METAL [25]. P-values were corrected for multiple testing using the Benjamini-Hochberg (FDR) correction [26]; resulting values are reported as *q-*values. CpG-sites were annotated using the manufacturer’s manifest (http://webdata.illumina.com.s3-website-us-east-1.amazonaws.com/downloads/productfiles/methylationEPIC/infinium-methylationepic-v-1-0-b4-manifest-file-csv.zip; downloaded on 10th of August 2018). Regression coefficients of differential methylation for the epigenome-wide significant CpG-sites were summarized for each brain region. As each brain region was processed on a separate plate, no inferential statistical procedure was applied to compare DNA-methylation levels between brain regions (due to confounding of batch and regions). Test statistics from all epigenome-wide significant CpG-sites were reported for each brain region and also for an independent EWAS in peripheral blood, in which DNA-methylation levels of male patients with AUD, who had just entered withdrawal treatment were compared with healthy controls [27]. As the cause of death of two control subjects from the second batch was “toxicity”, a sensitivity analysis excluding these subjects was performed in CN, VS, and PUT.

#### Differentially methylated regions (DMRs)

DMRs were identified using the comb-p algorithm [28], which accounts for autocorrelation between tests of adjacent methylation sites and combines these sites to regions of enrichment, in a given window. The following settings were used: Seed-p value < 0.01, minimum of 2 probes, sliding window 500 bp. The Šidák correction as implemented in comb-p was applied to correct for multiple testing. Comb-p was applied to the result statistics for all brain regions.

#### Pyrosequencing and TaqMan Assay

The DMR in *DDAH2* was replicated by pyrosequencing and gene expression levels were determined using a TaqMan Assay (for details see Supplementary Text S3).

#### Gene-Ontology (GO) over-representation analysis

Functional analysis to identify gene pathways targeted by differentially methylated CpG-sites was performed for sites with a threshold of *p*_*nominal*_ < 0.001 using missMethyl [29]. missMethyl controls for probe number bias, the increased likelihood of a gene to be differentially methylated, if more probes cover the gene and multi-gene bias, and the fact that probes can be annotated to more than one gene.

#### GWAS-Enrichment-Analysis

Gene-sets were created consisting of the genes to which the differentially methylated CpG-sites were annotated. Two gene-sets were created for each of the CN and VS results, one for genes implicated by epigenome-wide significant CpG-sites, and one for genes implicated by nominally significant CpG-sites, giving a total of four gene-sets. Multi-marker Analysis of GenoMic Annotation (MAGMA)[30] was used to test enrichment of those gene-sets in the results of a genome-wide association study (GWAS) of AUD [31].

#### Weighted Correlation Network Analysis (WGCNA)

The WGCNA R package [32] was used to generate co-methylated modules and relate those to AUD case-/control status. For each brain region the quantile-normalized beta values of CpG-sites nominally associated (*p* < 0.05) with AUD status were used as input. Soft power thresholds were picked according to the criterion of approximate scale-free topology (R_signed_^2^ > 0.90). The number of CpG-sites and the soft power thresholds picked can be found in Supplementary Table S2. Unassigned CpG-sites were clustered in the “grey” module, which was not taken into account for further analyses. For each brain region, the module of correlated CpG-sites with the highest association with AUD was identified. A GO analysis with the CpG-sites comprising the module was performed using missMethyl [29].

#### GWAS ATLAS

The PheWAS tool from the publicly available database GWAS ATLAS [33] [https://atlas.ctglab.nl/] was used to identify genome-wide significant associations of the genes implied by the top hits in the EWAS.

## Results

### Epigenome-wide association Analysis

In the CN, two CpG-sites were epigenome-wide significantly hypomethylated in AUD cases compared to controls. The two sites were annotated to the genes *IREB2* (cg04214706) and *HMGCR* (cg26685658). cg04214706 was also differentially methylated in the ACC (*p*_*nominal*_ = 0.005).

In the VS, 18 CpG-sites were epigenome-wide significantly associated with AUD. Nine CpG-sites were hyper- and nine hypomethylated. The top three hits were annotated to *SLC30A8, FAM20B*, and *PCAT29*. Of the epigenome-wide significant CpG-sites, cg12049992 in *PIEZO2* and cg16767842 in *GLANT9* were also differentially methylated in CN (*p*_*nominal*_ ≤ 0.023). Additionally, cg1354575 in *TCL1A* was differentially methylated in PUT (*p*_*nominal*_ = 0.035) and cg02849689 (intergenic) in ACC (*p*_*nominal*_ = 0.012). Three of the epigenome-wide significant CpG-sites showed nominally significant associations in an EWAS of AUD in peripheral blood, namely cg27512762 in *PCAT29*, cg06427508 in KLHL6 (effect in opposite direction), and cg02849689, which was not annotated to a nearby gene. In ACC, BA9, and PUT no epigenome-wide significant differentially methylated CpG-sites were identified (*q* ≥ 0.57). Epigenome-wide significant CpG-sites can be found in Table 3 and the top 100 associations for each brain region, together with more detailed information on location and annotation to enhancers, in supplementary tables (S3a–S3e). All coefficients of CpG-sites with *q <* 0.05 for each brain region and in peripheral blood are summarized in Supplementary Table S4. Manhattan plots for EWAS in the ACC, CN, and VS are depicted in Figure 1. Post-hoc power analyses using the web app EPIC Array Power Calculations (https://epigenetics.essex.ac.uk/shiny/EPICDNAmPowerCalcs/) were conducted for sample sizes of n=46 and n=94, to adequately reflect our sample sizes and the additional settings 2% mean difference and significance threshold 1×10^−7^, which was closest to the FDR corrected thresholds in the present study. This resulted in 11% of CpG-sites having a power larger than 90% to detect mean methylation differences of 2% for a sample size of 94 and 3.18% for a sample size of 46 (see also Supplementary Figure S1). It has to be noted, that the power calculations assumed equal distributions between cases and controls, which was not the case for all analyses. The sensitivity analyses did not reveal major differences between the EWAS in the complete sample and the reduced sample, in which control subjects who died of toxicity were excluded. The effect sizes of the nominally significant CpG-sites in each of the brain regions were highly correlated (*r*_CN_ = 0.99, *r*_VS_ = 0.98, *r*_PUT_ = 0.99, all *p* < 0.001). Scatterplots of the effect sizes for nominally significant CpG-sites in both analyses are depicted in Supplementary Figure S2.

**Table 3.**
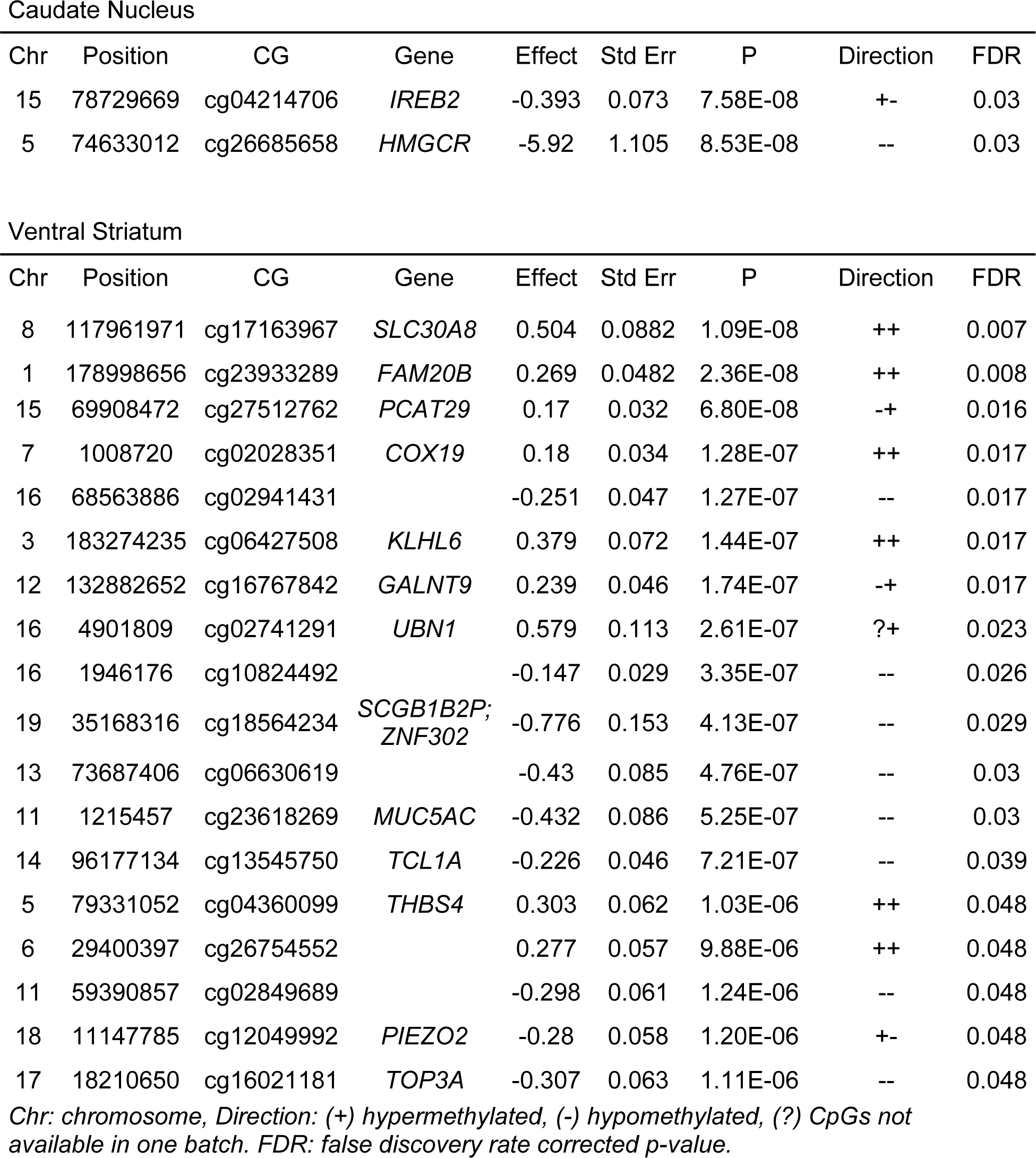
Epigenome-wide significant CpG-sites associated with AUD.

**Figure 1.**
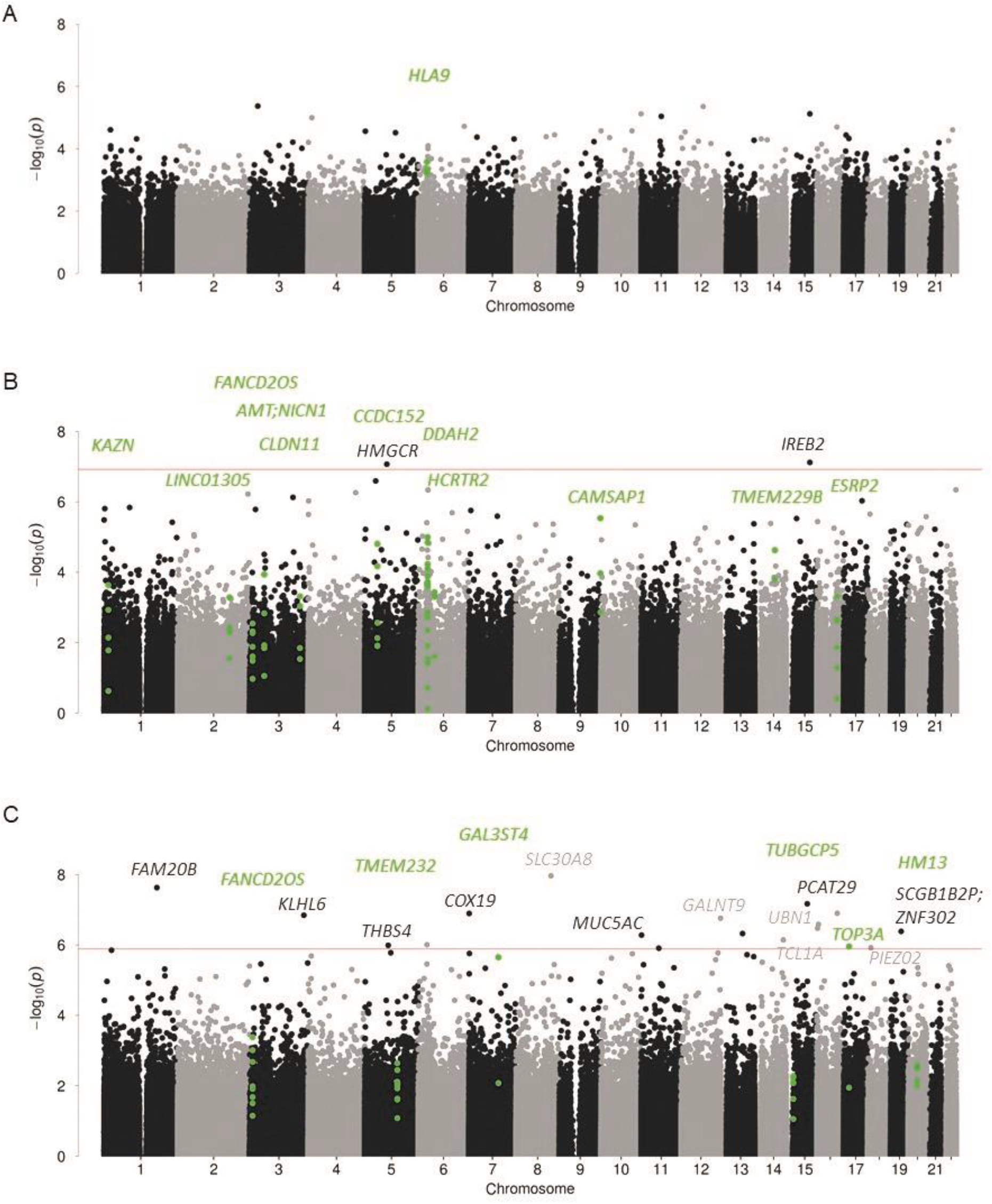
Manhattan plots of association of methylation values with AUD in (A) anterior cingulate cortex; (B) caudate nucleus; (C) ventral striatum. Highlighted CpG-sites represent differentially methylated regions. Genes implicated by CpGs (light and dark grey) and DMRs (green) are specified in the figures. Red line indicates FDR-corrected significance.

### Differentially methylated regions

In the CN, 10 DMRs were associated with AUD. The top three regions were annotated to the genes *DDAH2, CCDC152*, and *CAMSAP1*. Six DMRs were associated with AUD (*q* < 0.05) in the VS, with the three most strongly associated regions in *TMEM232, FANCD2OS*, and *HM13*. All significant DMRs for CN and VS are highlighted in figure 1 and can be found in supplementary tables S5a and S5b. In the ACC, one region in *HLA9* was differentially methylated (*p* _Šidák *-corrected*_ = 3.25*10^−6^). No epigenome-wide significant DMRs were observed in BA9 and PUT.

The DMR in *DDAH2* was replicated by pyrosequencing (cg04074004: *t*(76.77)= 2.39, *p* = 0.019). Differential expression was not observed (all *p* > 0.136). For details see Supplementary Text S3.

### Gene-Ontology Analysis

The strongest overrepresentation in the CN was for the biological process “homophilic cell adhesion via plasma membrane adhesion molecules” (*p* = 5.37*10^−6^, *q* = 0.12) and “cell-cell adhesion via plasma-membrane adhesion molecules” (*p* = 1.68*10^−5^, *q* = 0.187). In the VS, the cellular “Lsm1-7-Pat1 complex” showed the strongest overrepresentation (*p* = 6.49*10^−5^, *q* ≈ 1). Both associations did not remain significant after correction for multiple testing. The ten GO-terms showing the strongest overrepresentation can be found in Supplementary Tables S6a and S6b.

### GWAS enrichment analysis

No significant enrichment was observed in any of the regions and gene-sets tested (all *p* ≥ 0.277).

### Weighted Correlation Network Analysis (WGCNA)

For the caudate nucleus, 15 modules were identified consisting of 49-10,330 CpG-sites (*Median* = 965). The strongest association with AUD was observed for module “black”, which showed the strongest enrichment for the cellular component “PML body” (*p* = 0.001) and the molecular function “G-rich strand telomeric DNA binding” (*p* = 0.001). For CpG-sites nominally associated with AUD status in the VS 14 modules were identified, consisting of 38-12,721 CpG-sites (*Median* = 611). Module “purple” showed the strongest association with AUD and was enriched for a variety of immune-related GO-terms, such as the biological processes “regulation of T-cell proliferation” (*p* = 4.32e^-6^) and “regulation of leukocyte cell-cell adhesion” (*p* = 6.83e^-6^). For caudate nucleus module “black” and ventral striatum module “purple” the correlations of the gene significance (GS), which reflects the biological significance of a CpG-site with an external trait (here AUD) and the module membership (MM), which reflects the correlation of each CpG-site with the module, were calculated and are displayed in Figure 2a and 2b. The top enriched GO-terms for these modules can be found in Supplementary Tables S7a and S7b. Results for ACC, BA9, and putamen are described in the Supplementary Information (Text S2, Figure S3).

**Figure 2.**
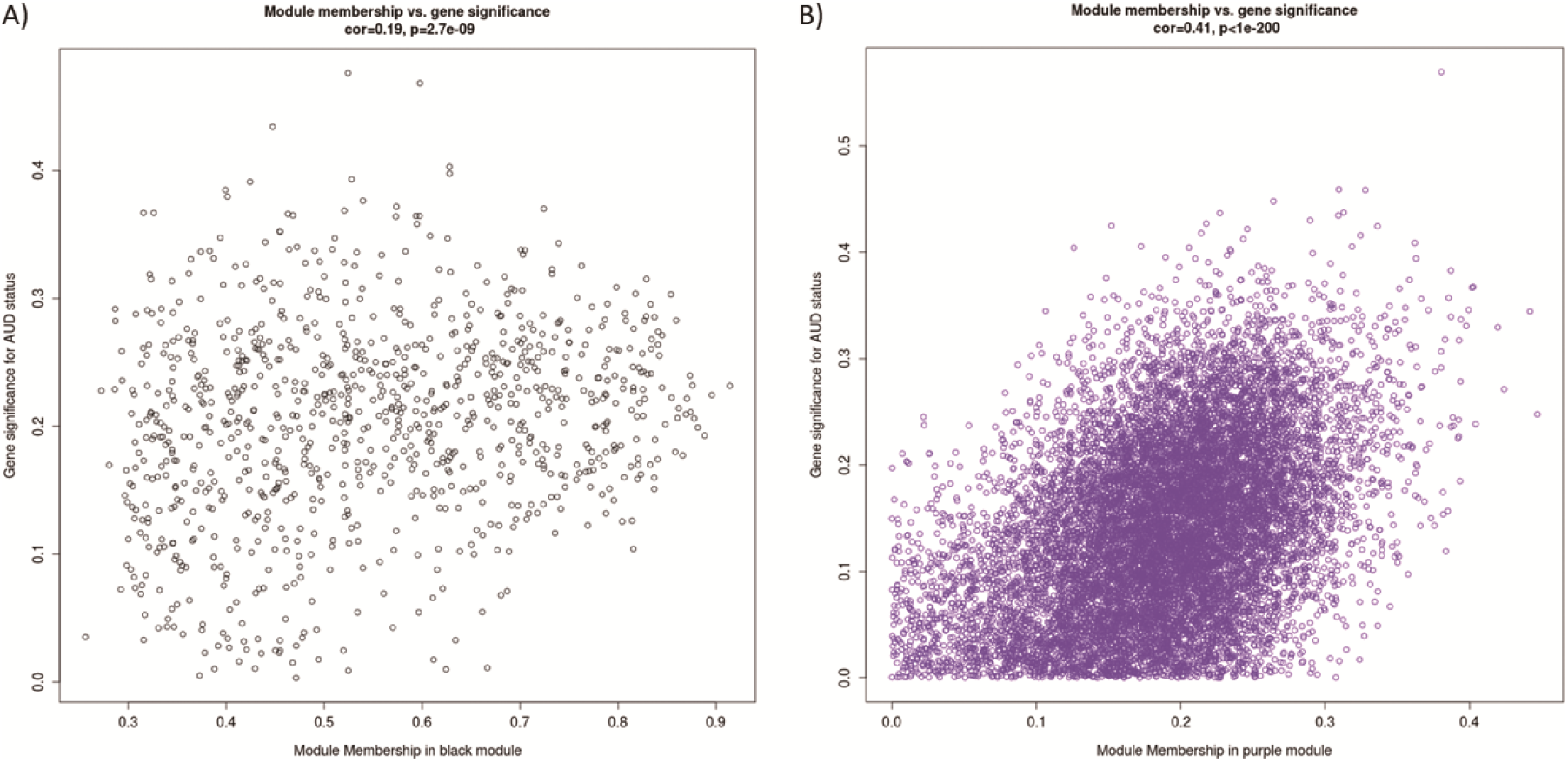
Association of gene significance for AUD status with module membership, for the modules A) “black” in caudate nucleus, and B) “purple” in ventral striatum.

### GWAS ATLAS

GWAS ATLAS results for the genes implicated by the most strongly associated site and region both in the CN and VS can be found in Supplementary Tables S8a-S8d. In brief, *IREB2* has previously been associated with smoking phenotypes (e.g., number of cigarettes a day, numbers of cigarettes previously smoked daily), parental illnesses such as lung cancer and chronic bronchitis, and psychiatric disorders like schizophrenia and bipolar disorder [33-35]. Genome-wide significant associations of *DDAH2* with phenotypes from a variety of domains, e.g., immunological, metabolic, respiratory, and psychiatric have been found. In the psychiatric domain, *DDAH2* has been associated with schizophrenia and bipolar disorder e.g. [34,36]. *SLC30A8* has been implied in blood sugar levels [37] and *TMEM232* in allergic rhinitis and asthma [33].

## Discussion

The present study examined DNA-methylation associated with AUD in regions of the addiction neurocircuitry using an epigenome-wide methylation analysis approach employed in human postmortem brain tissue. The largest of its kind to date and first to examine five brain regions, this study identified several novel differentially methylated CpG-sites as well as DMRs associated with AUD, providing potential insight into underlying mechanisms.

We found significant differentially methylated CpG-sites in two striatal regions. In the caudate nucleus, two epigenome-wide significant CpG-sites in *IREB2* and *HMGCR*, were identified. *IREB2* is a gene encoding iron regulatory protein 2, which is an RNA-binding protein that is involved in the regulation of cellular iron metabolism [https://www.genecards.org/cgi-bin/carddisp.pl?gene=IREB2]. Iron overload in the brain has previously been associated with cognitive decline in AUD [38]. Neurodegeneration has been reported in two subjects with bi-allelic loss of function variants in *IREB2* [39,40]. *IREB2* has also been associated with smoking phenotypes [33]. The association with smoking, which strongly affects DNA methylation [41,42], may be linked to the relevance of the gene to addiction phenotypes. In the present study, the *IREB2-*CpG-site was also differentially methylated in the anterior cingulate cortex (nominal significance), which might reflect a relevance in addiction phenotypes in multiple brain regions.

In the ventral striatum, 18 CpG-sites were epigenome-wide significantly associated with AUD. The strongest association was observed in a CpG-site in *SLC30A8*, which encodes a zinc efflux transporter that is involved in the accumulation of zinc in the intracellular vesicles. Zinc is a structure-building element in alcohol dehydrogenase (ADH) and thereby important for the proper function of ADH, which is needed to break down alcohol [43]. Differential methylation in *SCL30A8* may lead to altered Zinc availability and indirectly impact ADH function, and thus alcohol metabolism. *SLC30A8* has also been implicated in type 1 and type 2 diabetes [44]. In both types epigenetic and transcriptomic levels of SLC30A8 have shown to be altered [45]. Heavy alcohol consumption is also an established risk factor for type 2 diabetes on the phenotypic level [46]. Three of the epigenome-wide significant CpG-sites were also previously differentially methylated in an independent EWAS of AUD [27]; these convergent results might point towards a cross-tissue effect of these sites.

Significant regional methylation differences were observed in the anterior cingulate cortex, caudate nucleus, and ventral striatum. One differentially methylated region was observed in the anterior cingulate cortex and that region was annotated to HLA complex group 9, a non-coding RNA in the major histocompatibility complex (MHC). HLA antigens play a role in AUD and alcohol-associated liver disease [47]. In the caudate nucleus, the DMR showing the strongest association with AUD was annotated to *DDAH2*, encoding for dimethylarginine dimethylaminohydrolase, which is involved in the formation of nitric oxide by indirect inhibition of nitric oxide synthase (NOS) [https://www.genecards.org/cgi-bin/carddisp.pl?gene=DDAH2]. Nitric oxide has previously been associated with sleep disturbances, as part of the sleep-wake state controlling metabolites [48]. Sleep disorders and disturbances, such as decreased total sleep time and decreased sleep efficiency, are common in individuals during periods of alcohol consumption and prolonged withdrawal [49,50]. In rodent studies, alcohol exposure influenced NOS expression in the brain [51] and the knockout of neuronal NOS was associated with increased consumption of highly concentrated alcohol solutions [52]. Although the DMR in *DDAH2* was replicated by pyrosequencing, we did not observe differential gene expression between AUD cases and controls. It has to be noted, that differential gene expression is only partly explained by DNA-methylation differences. For example, in postmortem brain samples of individuals with schizophrenia and controls, 204 of 71,753 tested CpG gene pairs were significantly correlated [53]. A potential functional relevance of the DMR in *DDAH2* requires further investigation, for instance in relation to contact frequency maps (chromosomal architecture/Hi-C), which can be simultaneously studied with the methylome in single-cell experiments [54].

Of the six DMRs identified in the ventral striatum, a region in *TMEM232* showed the strongest association. *TMEM232* has previously been associated with respiratory traits, such as seasonal allergic rhinitis [55]. Another significant CpG-site was annotated to *HM13*. This gene encodes for minor histocompatibility antigen H13. In general, minor histocompatibility antigens function in the immune system by recognizing T cells [56]. No studies have investigated direct associations between AUD and H13 expression changes yet, but it is known that the immune system is downregulated in patients with AUD [57].

GO-term analyses investigating molecular functions associated with differentially methylated CpG-sites did not yield significant results after multiple testing correction, which is most likely attributable to the limited statistical power. No significant enrichment was observed for each of the gene-sets in GWAS signals for AUD, which could indicate that differential methylation in the newly identified CpG-sites is more sensitive to environmental factors than genetic effects.

In the WGCNA analysis in VS a module enriched for immune-processes was most strongly associated with AUD, which are known to be influenced by alcohol abuse [58].

In this brain region-specific analysis, comparing individuals with AUD and controls, we focused beside prefrontal areas on striatal regions, as previous studies have indicated that AUD may be associated with a striatal shift in activation from ventral to dorsal, as drug intake changes from goal-directed to habitual [59,60]. These studies focus on changes in neurotransmitter release and functional connectivity but it is not known how epigenetic changes impact this functional striatal shift. Our epigenome-wide results provide a first basis to explore epigenetic contributions to functional striatal changes.

This study has several limitations. The first is PMI, which can influence the tissue quality. The longer the individual has been deceased before the tissue was extracted from the body, the further along are degradation processes [61]. While we corrected for this in our analyses our results may have been affected by postmortem degradation processes nevertheless. Second, we cannot infer whether the observed differences in DNA-methylation are a result of addiction or long-term alcohol consumption, which affects multiple organ systems. Third, the methylation array used in the present study combined with the bisulfite conversion does not distinguish between methylation and hydroxymethylation. Therefore, no conclusions can be drawn regarding methylation type specific effects. Also, for several CpG-sites the effect in the meta-analysis was driven by a large effect in one, but not the other batch and in some of the cases this went hand-in-hand with a change in direction. For example, cg04214706 had a small positive effect, which was statistically not different from zero in the first batch, and a large negative effect in the second. Further samples are needed to validate these findings. Due to the sparse availability of human postmortem brain tissue, our sample size is small compared to EWAS in peripheral blood, which results in limited statistical power, especially taking into account the high multiple testing correction burden. However, EWAS analysis of peripheral blood allows to reveal only limited conclusions about differential methylation in the brain, whereas studies that examine multiple brain sites in a comparative fashion point to region-specific functional changes. Lastly, the correlational design of this analysis does not allow conclusions about the causality of the findings. DNA-methylation differences both be a result of AUD and be present in individuals before onset of the disorder.

Here, we identified novel associations of differential DNA-methylation between AUD cases and controls, which are prominent in alcohol-related pathways and diseases linked with AUD. To confirm these observations, larger samples are needed from the respective brain regions. Human postmortem brain tissue is difficult to obtain and very few brain banks focus on substance use disorders. Combining existing datasets, generating a larger amount of DNA-methylation data, and integrating multi-omics data, could lead to more conclusive results that may help to understand the molecular changes due to substance abuse in the brain and eventually to the identification of drug targets for more effective treatment of substance use disorders.

## Data Availability

Raw data and summary statistics for all analyses are available on request.

## Acknowledgments

We thank Elisabeth Röbel and Claudia Schäfer-Arnold for technical assistance.

Tissues were received from the New South Wales Brain Tissue Resource Centre at the University of Sydney which is supported by the University of Sydney. Research reported in this publication was supported by the National Institute of Alcohol Abuse and Alcoholism of the National Institutes of Health under Award Number R28AA012725. The content is solely the responsibility of the authors and does not represent the official views of the National Institutes of Health.

## Author Contributions

SHW, MR, RS, MMN, and ACH planned the investigation. MMF and ACH performed the DNA and RNA extraction and SHH, PH, FD and MMN were responsible for generating genome-wide methylation data. LZ, JF, JCF and FS developed the analysis plan. LZ and JF performed all statistical analyses. HD, ACH and MMF performed replication and validation experiments. LZ, MMF, JCF, LS, MR, FS, and SHW reviewed the literature for the paper. LZ, MMF, FS and SHW drafted the manuscript. All authors contributed, revised, and edited the final manuscript critically. All authors agreed to the publication of the final version of the manuscript.

## Data Availability

Raw data and summary statistics for all analyses are available on request.

## References

1 World Health Organization. Global status report on alcohol and health 2018. World Health Organization; 2019.

2 Degenhardt L, Charlson F, Ferrari A, Santomauro D, Erskine H, Mantilla-Herrara A, et al. The global burden of disease attributable to alcohol and drug use in 195 countries and territories, 1990–2016: a systematic analysis for the Global Burden of Disease Study 2016. The Lancet Psychiatry. 2018;5(12):987–1012.

3 Verhulst B, Neale MC, Kendler KS. The heritability of alcohol use disorders: a meta-analysis of twin and adoption studies. Psychological medicine. 2015;45(5):1061.

4 Robison AJ, Nestler EJ. Transcriptional and epigenetic mechanisms of addiction. Nat Rev Neurosci. 2011;12(11):623–37.

5 Longley MJ, Lee J, Jung J, Lohoff FW. Epigenetics of alcohol use disorder—A review of recent advances in DNA methylation profiling. Addict Biol. 2021:e13006.

6 Maze I, Nestler EJ. The epigenetic landscape of addiction. Ann N Y Acad Sci. 2011;1216:99–113.

7 Wedemeyer F, Kaminski JA, Zillich L, Hall ASM, Friedel E, Witt SH. Prospects of Genetics and Epigenetics of Alcohol Use Disorder. Current Addiction Reports. 2020.

8 Rakyan VK, Down TA, Balding DJ, Beck S. Epigenome-wide association studies for common human diseases. Nat Rev Genet. 2011;12(8):529–41.

9 Lohoff FW, Roy A, Jung J, Longley M, Rosoff DB, Luo A, et al. Epigenome-wide association study and multi-tissue replication of individuals with alcohol use disorder: evidence for abnormal glucocorticoid signaling pathway gene regulation. Molecular Psychiatry. 2020:1–14.

10 Edgar RD, Jones MJ, Meaney MJ, Turecki G, Kobor MS. BECon: a tool for interpreting DNA methylation findings from blood in the context of brain. Transl Psychiatry. 2017;7(8):e1187.

11 Goldstein RZ, Volkow ND. Dysfunction of the prefrontal cortex in addiction: neuroimaging findings and clinical implications. Nat Rev Neurosci. 2011;12(11):652–69.

12 Wang F, Xu H, Zhao H, Gelernter J, Zhang H. DNA co-methylation modules in postmortem prefrontal cortex tissues of European Australians with alcohol use disorders. Scientific reports. 2016;6:19430.

13 Gatta E, Grayson DR, Auta J, Saudagar V, Dong E, Chen Y, et al. Genome-wide methylation in alcohol use disorder subjects: implications for an epigenetic regulation of the cortico-limbic glucocorticoid receptors (NR3C1). Molecular Psychiatry. 2021;26(3):1029–41.

14 Koob GF, Volkow ND. Neurocircuitry of addiction. Neuropsychopharmacol. 2010;35(1):217–38.

15 Noori HR, Spanagel R, Hansson AC. Neurocircuitry for modeling drug effects. Addict Biol. 2012;17(5):827–64.

16 Park SQ, Kahnt T, Beck A, Cohen MX, Dolan RJ, Wrase J, et al. Prefrontal cortex fails to learn from reward prediction errors in alcohol dependence. J Neurosci. 2010;30(22):7749–53.

17 Volkow ND, Morales M. The brain on drugs: from reward to addiction. Cell. 2015;162(4):712–25.

18 Galandra C, Basso G, Cappa S, Canessa N. The alcoholic brain: neural bases of impaired reward-based decision-making in alcohol use disorders. Neurol Sci. 2018;39(3):423–35.

19 Meng W, Sjöholm LK, Kononenko O, Tay N, Zhang D, Sarkisyan D, et al. Genotype-dependent epigenetic regulation of DLGAP2 in alcohol use and dependence. Molecular Psychiatry. 2019:1–16.

20 Lehne B, Drong AW, Loh M, Zhang W, Scott WR, Tan S-T, et al. A coherent approach for analysis of the Illumina HumanMethylation450 BeadChip improves data quality and performance in epigenome-wide association studies. Genome biology. 2015;16(1):37.

21 Du P, Zhang X, Huang C-C, Jafari N, Kibbe WA, Hou L, et al. Comparison of Beta-value and M-value methods for quantifying methylation levels by microarray analysis. BMC bioinformatics. 2010;11(1):587.

22 Houseman EA, Accomando WP, Koestler DC, Christensen BC, Marsit CJ, Nelson HH, et al. DNA methylation arrays as surrogate measures of cell mixture distribution. BMC Bioinformatics. 2012;13(1):86.

23 Jaffe AE, Kaminsky ZA. (R package version 1.24.0, 2020).

24 Maas SC, Vidaki A, Wilson R, Teumer A, Liu F, van Meurs JB, et al. Validated inference of smoking habits from blood with a finite DNA methylation marker set. European journal of epidemiology. 2019;34(11):1055–74.

25 Willer CJ, Li Y, Abecasis GR. METAL: fast and efficient meta-analysis of genomewide association scans. Bioinformatics. 2010;26(17):2190–91.

26 Benjamini Y, Hochberg Y. Controlling the false discovery rate: a practical and powerful approach to multiple testing. Journal of the Royal Statistical Society: series B. 1995;57(1):289–300.

27 Witt SH, Frank J, Frischknecht U, Treutlein J, Streit F, Foo JC, et al. Acute alcohol withdrawal and recovery in men lead to profound changes in DNA methylation profiles: a longitudinal clinical study. Addiction. 2020.

28 Pedersen BS, Schwartz DA, Yang IV, Kechris KJ. Comb-p: software for combining, analyzing, grouping and correcting spatially correlated P-values. Bioinformatics. 2012;28(22):2986–88.

29 Phipson B, Maksimovic J, Oshlack A. missMethyl: an R package for analyzing data from Illumina’s HumanMethylation450 platform. Bioinformatics. 2016;32(2):286–88.

30 de Leeuw CA, Mooij JM, Heskes T, Posthuma D. MAGMA: Generalized Gene-Set Analysis of GWAS Data. PLOS Computational Biology. 2015;11(4):e1004219.

31 Zhou H, Sealock JM, Sanchez-Roige S, Clarke T-K, Levey DF, Cheng Z, et al. Genome-wide meta-analysis of problematic alcohol use in 435,563 individuals yields insights into biology and relationships with other traits. Nature Neuroscience. 2020:1–10.

32 Langfelder P, Horvath S. WGCNA: an R package for weighted correlation network analysis. BMC bioinformatics. 2008;9(1):559.

33 Watanabe K, Stringer S, Frei O, Umicevic Mirkov M, de Leeuw C, Polderman TJC, et al. A global overview of pleiotropy and genetic architecture in complex traits. Nat Genet. 2019;51(9):1339–48.

34 Ruderfer DM, Ripke S, McQuillin A, Boocock J, Stahl EA, Pavlides JMW, et al. Genomic dissection of bipolar disorder and schizophrenia, including 28 subphenotypes. Cell. 2018;173(7):1705-15. e16.

35 Furberg H, Kim Y, Dackor J, Boerwinkle E, Franceschini N, Ardissino D, et al. Genome-wide meta-analyses identify multiple loci associated with smoking behavior. Nat Genet. 2010;42(5):441.

36 Pardinas AF, Holmans P, Pocklington AJ, Escott-Price V, Ripke S, Carrera N, et al. Common schizophrenia alleles are enriched in mutation-intolerant genes and in regions under strong background selection. Nat Genet. 2018;50(3):381–89.

37 Kanai M, Akiyama M, Takahashi A, Matoba N, Momozawa Y, Ikeda M, et al. Genetic analysis of quantitative traits in the Japanese population links cell types to complex human diseases. Nat Genet. 2018;50(3):390–400.

38 Listabarth S, König D, Vyssoki B, Hametner S. Does thiamine protect the brain from iron overload and alcohol-related dementia? Alzheimers Dement. 2020;16(11):1591–95.

39 Costain G, Ghosh MC, Maio N, Carnevale A, Si YC, Rouault TA, et al. Absence of iron-responsive element-binding protein 2 causes a novel neurodegenerative syndrome. Brain. 2019;142(5):1195–202.

40 Cooper MS, Stark Z, Lunke S, Zhao T, Amor DJ. IREB2-associated neurodegeneration. Brain. 2019;142(8):e40.

41 Zeilinger S, Kühnel B, Klopp N, Baurecht H, Kleinschmidt A, Gieger C, et al. Tobacco smoking leads to extensive genome-wide changes in DNA methylation. PloS one. 2013;8(5):e63812.

42 Sugden K, Hannon EJ, Arseneault L, Belsky DW, Broadbent JM, Corcoran DL, et al. Establishing a generalized polyepigenetic biomarker for tobacco smoking. Transl Psychiatry. 2019;9(1):92.

43 Baj J, Flieger W, Teresinski G, Buszewicz G, Sitarz E, Forma A, et al. Magnesium, Calcium, Potassium, Sodium, Phosphorus, Selenium, Zinc, and Chromium Levels in Alcohol Use Disorder: A Review. J Clin Med. 2020;9(6):1901.

44 Sladek R, Rocheleau G, Rung J, Dina C, Shen L, Serre D, et al. A genome-wide association study identifies novel risk loci for type 2 diabetes. Nature. 2007;445(7130):881–5.

45 Gu HF. Genetic, epigenetic and biological effects of zinc transporter (SLC30A8) in type 1 and type 2 diabetes. Curr Diabetes Rev. 2017;13(2):132–40.

46 Baliunas DO, Taylor BJ, Irving H, Roerecke M, Patra J, Mohapatra S, et al. Alcohol as a risk factor for type 2 diabetes: a systematic review and meta-analysis. Diabetes Care. 2009;32(11):2123–32.

47 Shigeta Y, Ishii H, Takagi S, Yoshitake Y, Hirano T, Takata H, et al. HLA antigens as immunogenetic markers of alcoholism and alcoholic liver disease. Pharmacol Biochem Behav. 1980;13:89–94.

48 Cespuglio R, Amrouni D, Meiller A, Buguet A, Gautier-Sauvigne S. Nitric oxide in the regulation of the sleep-wake states. Sleep Med Rev. 2012;16(3):265–79.

49 Thakkar MM, Sharma R, Sahota P. Alcohol disrupts sleep homeostasis. Alcohol. 2015;49(4):299–310.

50 Koob GF, Colrain IM. Alcohol use disorder and sleep disturbances: a feed-forward allostatic framework. Neuropsychopharmacol. 2020;45(1):141–65.

51 Davis RL, Syapin PJ. Interactions of alcohol and nitric-oxide synthase in the brain. Brain Res Brain Res Rev. 2005;49(3):494–504.

52 Spanagel R, Siegmund S, Cowen M, Schroff K-C, Schumann G, Fiserova M, et al. The neuronal nitric oxide synthase gene is critically involved in neurobehavioral effects of alcohol. J Neurosci. 2002;22(19):8676–83.

53 Chen C, Zhang C, Cheng L, Reilly JL, Bishop JR, Sweeney JA, et al. Correlation between DNA methylation and gene expression in the brains of patients with bipolar disorder and schizophrenia. Bipolar Disord. 2014;16(8):790–99.

54 Li G, Liu Y, Zhang Y, Kubo N, Yu M, Fang R, et al. Joint profiling of DNA methylation and chromatin architecture in single cells. Nature Methods. 2019;16(10):991–93.

55 Ramasamy A, Curjuric I, Coin LJ, Kumar A, McArdle WL, Imboden M, et al. A genome-wide meta-analysis of genetic variants associated with allergic rhinitis and grass sensitization and their interaction with birth order. J Allergy Clin Immunol 2011;128(5):996–1005.

56 Robertson NJ, Chai JG, Millrain M, Scott D, Hashim F, Manktelow E, et al. Natural regulation of immunity to minor histocompatibility antigens. J Immunol. 2007;178(6):3558–65.

57 Erickson EK, Grantham EK, Warden AS, Harris RA. Neuroimmune signaling in alcohol use disorder. Pharmacol Biochem Behav. 2019;177:34–60.

58 Pasala S, Barr T, Messaoudi I. Impact of Alcohol Abuse on the Adaptive Immune System. Alcohol research : current reviews. 2015;37(2):185–97.

59 Vollstädt-Klein S, Wichert S, Rabinstein J, Bühler M, Klein O, Ende G, et al. Initial, habitual and compulsive alcohol use is characterized by a shift of cue processing from ventral to dorsal striatum. Addiction. 2010;105(10):1741–49.

60 DePoy L, Daut R, Brigman JL, MacPherson K, Crowley N, Gunduz-Cinar O, et al. Chronic alcohol produces neuroadaptations to prime dorsal striatal learning. Proc Natl Acad Sci U S A. 2013;110(36):14783–8.

61 Glausier JR, Konanur A, Lewis DA. Factors Affecting Ultrastructural Quality in the Prefrontal Cortex of the Postmortem Human Brain. J Histochem Cytochem. 2019;67(3):185–202.

62 Heinz A, Kiefer F, Smolka MN, Endrass T, Beste C, Beck A, et al. Addiction Research Consortium: Losing and regaining control over drug intake (ReCoDe)—From trajectories to mechanisms and interventions. Addict Biol. 2020;25(2):e12866.

