## Supplemental Material for "Epigenome-wide Association Study of Alcohol Use Disorder in Five Brain Regions"

### Supplementary Information

Lea Zillich[1] (MSc), Josef Frank[1] (PhD), Fabian Streit[1] (PhD), Marion M Friske [2] (MSc), Jerome C Foo [1] (PhD), Lea Sirignano [1] (MSc), Stefanie Heilmann-Heimbach [3] (PhD), Helene Dukal (MSc)[1], Franziska Degenhardt [3,4] (MD), Per Hoffmann [3] (PhD), Anita C Hansson [2] (PhD), Markus M Nöthen [3] (MD), Marcella Rietschel[1] (MD), Rainer Spanagel [2]* (PhD), Stephanie H Witt[1,5]* (PhD)

*shared senior authorship

**Text S1.** Detailled description of obtained postmortem human brain samples.

**Text S2**. WGCNA results for Brodmann Area 9, Anterior Cingulate Cortex and Putamen.

**Text S3.** Methods and results for replication by pyrosequencing and functional validation for *DDAH2*.

**Table S1.** Additional Phenotype information.

**Table S2.** WGCNA input

**Tables S3a-3e.** Top 100 CpG-sites associated with AUD are provided in the Excel file for all brain regions.

**Table S4**. Regression coefficients for the epigenome-wide significant CpG-sites in other brain regions and in peripheral blood.

**Table S5a.** Results of comparison between AUD cases and controls (epigenome-wide significant regions) in the caudate nucleus.

**Table S5b.** Results of comparison between AUD cases and controls (epigenome-wide significant regions) in the ventral striatum.

**Table S6a.** Top 10 associated GO-terms for the AUD case/control comparison in the caudate nulceus.

**Table S6b.** Top 10 associated GO-terms for the AUD case/control comparison in the ventral striatum.

**Table S7a.** Top 10 associated GO-terms for the WGCNA-module "black" (caudate nucleus).

**Table S7b.** Top 10 associated GO-terms for the WGCNA-module "purple" (ventral striatum).

**Table S8a.** GWAS ATLAS results for *IREB2* [retrieved 12.01.2021].

**Table S8b.** GWAS ATLAS results for *SLC30A8* [retrieved 12.01.2021].

**Table S8c.** GWAS ATLAS results for *DDAH2* [retrieved 20.01.2021].

**Table S8d.** GWAS ATLAS results for *TMEM232* [retrieved 12.01.2021].

**Figure S1.** Scatterplots of effects for nominally associated CpG-sites from the EWAS in all samples, and from the sensitivity analysis for which subjects with blood alcohol at death were excluded.

**Figure S2**. Scatterplots of CpG-site significance and module membership, of WGCNA modules most strongly associated with AUD in anterior cingulate cortex, Brodmann Area 9, and Putamen.

**Figure S3**. Power curve showing the proportion of CpG-sites on the EPIC array with sufficient power to detect a mean methylation difference of 2%.

**Text S1.** Detailled description of obtained postmortem human brain samples.

All tissue samples were obtained from the New South Wales Tissue Resource Centre (University of Sydney, Australia). Tissues were collected as described in Harper et al. (2003, Progress in Neuro-Psychopharmacology & Biological Psychiatry 27: 951– 961), where cases with prolonged agonal life support, cases with a history of cerebral infarction, head injury, or neurodegenerative disease (e.g., Alzheimer's disease) were excluded. Further exclusion criteria were cases with developmental disorder, head injury at time of death, recent cerebral stroke, neurological disorder, history of intravenous drug use, brain on gross examination showing obvious abnormalities, and cases with a history of other psychiatric disorders (e.g. autism, attention deficit-hyperactivity disorder, schizophrenia). We further excluded cases with reported anxiety or mood disorders or suicides have been excluded as well as cases with significant amounts of psychiatric medication (e.g., opioids, benzodiazepines; concentration <0.1 mg/L).

**Text S2.** WGCNA results for Anterior Cingulate Cortex, Brodmann Area 9, and Putamen.

Weighted correlation network analysis for Anterior Cingulate Cortex resulted in 10 modules consisting of 34–11,588 CpG-sites (*Md* = 606.5). The highest correlation with AUD was observed for module "green" ($r$ = .44, $p$ = 8.54e-04), which was enriched for the molecular functions "ligand-gated sodium channel activity" ($p$ = 0.0003), "sodium channel activity" ($p$ = 0.0004), and the biological process "noradrenergic neuron differentiation" ($p$ = 0.0007). In Brodmann Area 9 20 modules, with a median sizes of 287 CpG-sites per module (range: 38-7,960) were constructed. Module "blue" showed the highest correlation with AUD ($r$ = -.55, $p = 4e^{-05}$). GO-analysis revealed enrichment for the cellular components "actomyosin", ($p$ = 0.0001), "stress fiber" ($p$ = 0.0007), and "contractile actin filament bundle" ($p$ = 0.0007). Eight modules were constructed for putamen, which consisted of 78-13,091 CpG-site with a median size of 2,020. Module "yellow" had the highest correlation with AUD ($r$ = -.52, $p = 8.13e^{-08}$). This module was enriched for the biological processes "response to immune response of other organism involved in symbiotic interaction", ($p$ = 0.0004), "response to host immune response" ($p$ = 0.0004), and "ventral spinal cord interneuron fate commitment" ($p$ = 0.001). Scatterplots showing the association of gene significance and module membership for the three modules can be found in Supplementary Figure S2.

**Text S3.** Methods and results for replication by pyrosequencing and functional validation for *DDAH2*.

**Samples.** For a subsample of n=87 sufficient material for Pyrosequencing and the TaqMan Assay was available.

**Pyrosequencing**. The CpG site cg04074004 was analyzed by pyrosequencing, using the commercially available PyroMark CpG assay Hs_DDAH2_03_PM (Qiagen, Hilden, Germany). All samples (n=87) were carefully randomized on positions on sequencing plates. DNA (500ng) was bisulfite-treated using the EpiTect Fast 96 Kit (Qiagen, Hilden, Germany). We controlled for bisulfite conversion reaction, PCR amplification, and pyrosequencing reaction by including unmethylated and methylated DNA (EpiTect PCR Control DNA Set, Qiagen, Hilden, Germany).

The DNA fragment was amplified by PCR (PyroMark PCR Kit, Qiagen, Hilden, Germany) from 2 µl of bisulfite-treated DNA in a PCR volume of 25 µl. Successful amplification and the specificity of the PCR product (137 base pairs) was checked on an agarose gel. To ensure equal amplification in the CpG assay an additional mixture of DNA with a 50% DNA methylation level was prepared from unmethylated, converted and methylated, converted DNA (EpiTect PCR Control DNA Set, Qiagen, Hilden, Germany). Then, 8 µL of PCR product was processed for pyrosequencing analysis with following manufacturer´s recommendations (PyroMark Gold Q24 reagents, Qiagen, Hilden, Germany), sequenced with the sequencing primer Hs_DDAH2_03_PM (Qiagen, Hilden, Germany) with the PyroMark Q24 instrument (Qiagen, Hilden, Germany). The percentage of methylation was quantified using the PyroMark Q24 software version 2.0.8 Build 3 (Qiagen, Hilden, Germany). The sequencing was performed in triplicates.

**TaqMan Assay.** RNA was extracted from caudate nucleus samples using the RNeay microKit (Qiagen, Hilden, Germany). The analysis was performed on triplicates of each gene and sample using the PCR Master Mix and FAM dye-labeled TaqMan MGB probes from ThermoFisher Scientific (Massachusetts, US). The following primers have been used for the human DDAH2 gene: Forward primer sequence 1: 5-CAAAAGGCTGTCCGGGCAATGGCAG-3, position 854 on cDNA, RefSeq NM-013974.3, TaqMan assay ID Hs00967863_g1, 60 bp amplicon; forward primer sequence 2: 5- GCTCGTAGGCCAGAGGTCGATGGAG-3, position 560 on cDNA, RefSeq NM-013974.3, TaqMan assay ID Hs00967860_g1, 81 bp amplicon). As housekeeping genes the human 18S RNA (TaqMan assay ID Hs99999901_s1, 187 bp amplicon) and human Gapdh (TaqMan assay ID Hs02786624_g1, 157 bp amplicon) were used. SDS 2.2.2 software (ABI) was employed to analyze fluorescence intensity and calculation of the theoretical cycle number when a defined threshold was reached (Ct-value).

**Statistical Analysis.** R version 3.6.1 was used for quality control filtering and statistical analyses of the pyrosequencing results (http://www.r-project.org). We removed measurements marked as unreliable by the PyroMark software. Triplicate measurements were averaged after the removal of outliers (values deviating more than 3% for Pyrosequencing and more than 5% for TaqMan Assay). For expression of DDAH2, delta scores between the primer pairs and the housekeeping genes were calculated. We calculated student's t-Tests to compare DNA methylation of cg04074004 and delta-scores of gene expression between AUD cases and controls, as well as an ANCOVA with the same covariates as in the EWAS.

**Table S1.** Additional Phenotype information.

| Characteristic | Cases | Controls | *p* |
|---|---|---|---|
| Cell counts by region | | | |
| Anterior Cingulate Cortex | 0.30 (0.17) | 0.27 (0.17) | 0.584 |
| Brodmann Area 9 | 0.38 (0.05) | 0.40 (0.10) | 0.381 |
| Putamen | 0.21 (0.06) | 0.21 (0.05) | 0.857 |
| Caudate Nucleus | 0.23 (0.05) | 0.23 (0.05) | 0.84 |
| Ventral Striatum | 0.26 (0.07) | 0.25 (0.07) | 0.37 |
| Cause of death | | | |
| Cardiac | 14 | 23 | |
| Cardiovascular | 4 | 18 | |
| Vascular | 2 | 2 | |
| Hepatic | 6 | | |
| Infection | 5 | 1 | |
| COPD | 1 | 1 | |
| Cardiovascular / Respiratory | 1 | | |
| Respiratory | 1 | 1 | |
| Toxicity | 6 | 2 | |
| Hepatic/Infection | 1 | | |
| Blood loss | 1 | | |
| Pancreatic | 1 | | |
| Carcinoma | | 1 | |
| Suicide | | 1 | |
| Missing | 10 | 8 | |

*Data are presented as count (n) or mean (±SD), p: p-value of t-Test comparing cases and controls.*

**Table S2.** WGCNA input

| Brain Region | # CpG-Sites | Soft Power Threshold |
|---|---|---|
| ACC | 29059 | 3 |
| BA9 | 24157 | 5 |
| Caudate Nucleus | 34943 | 4 |
| Ventral Striatum | 35007 | 4 |
| Putamen | 28624 | 2 |

**Table S4**. Regression coefficients for the epigenome-wide significant CpG-sites in other brain regions and in peripheral blood.

| | Anterior Cingulate Cortex | | Brodmann Area 9 | | Putamen | | Caudate Nucleus | | Ventral Striatum | | Peripheral Blood | |
|---|---|---|---|---|---|---|---|---|---|---|---|---|
| CpG-Site | Effect | P | Effect | P | Effect | P | Effect | P | Effect | P | Effect | P |
| cg02028351 | 0.061 | 0.635 | -0.077 | 0.627 | -0.088 | 0.165 | -0.036 | 0.594 | 0.18 | **1.28E-07** | 0.012 | 0.808 |
| cg02741291 | | | | | 0.046 | 0.567 | 0.02 | 0.901 | 0.579 | **2.61E-07** | 0.014 | 0.860 |
| cg02849689 | 0.387 | **0.012** | -0.029 | 0.877 | 0.043 | 0.542 | -0.161 | 0.174 | -0.298 | **1.24E-06** | -0.070 | **0.045** |
| cg02941431 | -0.016 | 0.894 | -0.128 | 0.368 | -0.015 | 0.729 | -0.056 | 0.383 | -0.251 | **1.27E-07** | 0.057 | 0.232 |
| cg04214706 | -0.381 | **0.005** | 0.172 | 0.4 | -0.064 | 0.434 | -0.393 | **7.58E-08** | -0.093 | 0.348 | -0.139 | 0.082 |
| cg04360099 | -0.061 | 0.648 | 0.036 | 0.874 | 0.037 | 0.706 | 0.078 | 0.408 | 0.303 | **1.03E-06** | 0.096 | 0.183 |
| cg06427508 | -0.054 | 0.74 | 0.081 | 0.685 | -0.016 | 0.777 | 0.025 | 0.683 | 0.379 | **1.44E-07** | -0.111 | **0.024** |
| cg06630619 | 0.166 | 0.465 | 0.064 | 0.79 | -0.181 | 0.101 | 0.024 | 0.847 | -0.43 | **4.76E-07** | -0.041 | 0.525 |
| cg10824492 | -0.084 | 0.354 | -0.227 | 0.148 | 0.02 | 0.62 | -0.084 | 0.123 | -0.147 | **3.35E-07** | -0.025 | 0.662 |
| cg12049992 | 0.327 | 0.121 | 0.166 | 0.653 | -0.051 | 0.409 | -0.19 | **0.023** | -0.28 | **1.20E-06** | 0.085 | 0.296 |
| cg13545750 | 0.045 | 0.654 | 0.006 | 0.958 | -0.104 | **0.035** | -0.011 | 0.878 | -0.226 | **7.21E-07** | -0.018 | 0.628 |
| cg16021181 | 0.053 | 0.635 | 0.259 | 0.248 | 0.155 | 0.15 | 0.01 | 0.926 | -0.307 | **1.11E-06** | -0.023 | 0.822 |
| cg16767842 | 0.073 | 0.537 | 0.022 | 0.893 | 0.079 | 0.194 | 0.166 | **0.02** | 0.239 | **1.74E-07** | 0.054 | 0.623 |
| cg17163967 | -0.014 | 0.93 | 0.03 | 0.882 | 0.077 | 0.221 | -0.114 | 0.171 | 0.504 | **1.09E-08** | 0.034 | 0.693 |
| cg18564234 | -0.187 | 0.276 | 0.242 | 0.116 | -0.076 | 0.33 | -0.006 | 0.966 | -0.776 | **4.13E-07** | 0.012 | 0.851 |
| cg23618269 | -0.147 | 0.617 | -0.104 | 0.657 | -0.04 | 0.704 | 0.011 | 0.936 | -0.432 | **5.25E-07** | -0.099 | 0.349 |
| cg23933289 | 0.13 | 0.313 | -0.067 | 0.696 | -0.024 | 0.708 | -0.043 | 0.564 | 0.269 | **2.36E-08** | -0.042 | 0.455 |
| cg26685658 | 0.891 | 0.626 | 0.976 | 0.381 | -1.943 | 0.089 | -5.919 | **8.53E-08** | 0.632 | 0.629 | -0.392 | 0.263 |
| cg26754552 | -0.144 | 0.394 | -0.079 | 0.589 | -0.044 | 0.543 | 0.05 | 0.591 | 0.277 | **9.88E-07** | -0.063 | 0.173 |
| cg27512762 | -0.085 | 0.358 | -0.072 | 0.568 | -0.032 | 0.518 | 0.088 | 0.065 | 0.17 | **6.80E-08** | 0.115 | **0.002** |

**Table S5a.** Results of comparison between AUD cases and controls (epigenome-wide significant regions) in the caudate nucleus.

| Chr | Start | End | Min_p | N_probes | z_p | z_sidak_p | CG | Gene |
|---|---|---|---|---|---|---|---|---|
| 6 | 31696063 | 31696520 | 1.12E-17 | 17 | 1.85E-20 | 2.81E-17 | cg08550588 | *DDAH2* |
| 5 | 42756786 | 42757024 | 9.81E-07 | 6 | 3.11E-11 | 9.08E-08 | cg18472410 | *CCDC152* |
| 9 | 138800324 | 138800391 | 8.93E-06 | 3 | 2.83E-10 | 2.93E-06 | cg00022024 | *CAMSAP1* |
| 6 | 55039232 | 55039383 | 0.0002647 | 4 | 1.14E-08 | 5.26E-05 | cg10725720 | *HCRTR2* |
| 3 | 10149974 | 10150225 | 0.0005157 | 7 | 1.56E-07 | 0.0004319 | cg00166722 | *FANCD2OS* |
| 3 | 49459909 | 49460058 | 1.85E-05 | 5 | 1.57E-07 | 0.0007315 | cg21926782 | *AMT;NICN1* |
| 3 | 170136766 | 170136921 | 0.005362 | 4 | 4.40E-07 | 0.00197 | cg02241055 | *CLDN11* |
| 2 | 175190675 | 175190822 | 0.005362 | 4 | 5.10E-07 | 0.002406 | cg02216981 | *LINC01305* |
| 1 | 15272238 | 15272384 | 0.00151 | 5 | 5.86E-07 | 0.002783 | cg11648522 | *KAZN* |
| 14 | 67940414 | 67940426 | 0.001105 | 2 | 5.73E-08 | 0.003311 | cg06215536 | *TMEM229B* |
| 16 | 68270252 | 68270388 | 0.005186 | 6 | 7.70E-07 | 0.003925 | cg04513006 | *ESRP2* |

**Table S5b.** Results of comparison between AUD cases and controls (epigenome-wide significant regions) in the ventral striatum.

| Chr | Start | End | Min_p | N_probes | z_p | z_sidak_p | CG | Gene |
|---|---|---|---|---|---|---|---|---|
| 5 | 110062343 | 110062730 | 0.0001288 | 11 | 4.04E-11 | 7.25E-08 | cg23279021 | TMEM232 |
| 3 | 10149963 | 10150225 | 0.0001288 | 8 | 8.40E-10 | 2.23E-06 | cg10729496 | FANCD2OS |
| 20 | 30135144 | 30135292 | 0.0003445 | 5 | 3.61E-08 | 0.0001692 | cg15815607 | HM13 |
| 15 | 22833149 | 22833309 | 0.003964 | 6 | 5.16E-07 | 0.002237 | cg07959104 | TUBGCP5 |
| 17 | 18210638 | 18210651 | 0.009594 | 2 | 6.08E-07 | 0.03195 | cg03540694 | TOP3A |
| 7 | 99765805 | 99765819 | 0.00969 | 2 | 6.81E-07 | 0.03323 | cg18810646 | GAL3ST4 |

**Table S6a.** Top 10 associated GO-terms for the AUD case/control comparison in the caudate nulceus.

| Term | Ont | N | DE | P.DE | FDR |
|---|---|---|---|---|---|
| homophilic cell adhesion via plasma membrane adhesion molecules | BP | 153 | 28 | 5.37E-06 | 0.1197854 |
| cell-cell adhesion via plasma-membrane adhesion molecules | BP | 234 | 34 | 1.68E-05 | 0.1869303 |
| dodecenoyl-CoA delta-isomerase activity | MF | 3 | 3 | 6.74E-05 | 0.5008741 |
| carbon-oxygen lyase activity | MF | 67 | 12 | 0.0001516 | 0.6269051 |
| protein localization to chromosome | BP | 74 | 13 | 0.000175 | 0.6269051 |
| hydro-lyase activity | MF | 50 | 10 | 0.0001764 | 0.6269051 |
| protein localization to chromatin | BP | 22 | 7 | 0.0002047 | 0.6269051 |
| peptidyl-lysine dimethylation | BP | 15 | 6 | 0.000225 | 0.6269051 |
| establishment of protein localization to chromatin | BP | 8 | 4 | 0.0003527 | 0.8735079 |
| regulation of dendrite development | BP | 126 | 21 | 0.0005592 | 1 |

**Table S6b.** Top 10 associated GO-terms for the AUD case/control comparison in the ventral striatum.

| Term | Ont | N | DE | P.DE | FDR |
|---|---|---|---|---|---|
| Lsm1-7-Pat1 complex | CC | 5 | 4 | 6.49E-05 | 1 |
| T cell proliferation | BP | 167 | 20 | 0.0001892 | 1 |
| receptor signaling complex scaffold activity | MF | 24 | 8 | 0.0003144 | 1 |
| negative regulation of transcription from RNA polymerase II promoter in response to stress | BP | 12 | 5 | 0.0003586 | 1 |
| regulation of T cell proliferation | BP | 144 | 17 | 0.0003979 | 1 |
| organelle part | CC | 8811 | 553 | 0.0004222 | 1 |
| AP-1 adaptor complex | CC | 7 | 4 | 0.000459 | 1 |
| negative regulation of endoplasmic reticulum unfolded protein response | BP | 12 | 5 | 0.0005909 | 1 |
| positive regulation of male gonad development | BP | 7 | 4 | 0.0006815 | 1 |
| RNA polymerase binding | MF | 70 | 11 | 0.0008278 | 1 |

**Table S7a.** Top 10 associated GO-terms for the WGCNA-module “black” (caudate nucleus).

| Term | Ont | N | DE | P.DE | FDR |
|---|---|---|---|---|---|
| PML body | CC | 70 | 12 | .0012 | 1 |
| G-rich strand telomeric DNA binding | MF | 6 | 3 | .0014 | 1 |
| sperm mitochondrion organization | BP | 2 | 2 | .0017 | 1 |
| presynapse assembly | BP | 26 | 7 | .0017 | 1 |
| ventricular system development | BP | 20 | 6 | .0017 | 1 |
| cardiac ventricle development | BP | 96 | 15 | .0017 | 1 |
| ventricular septum development | BP | 55 | 11 | .0018 | 1 |
| type I transforming growth factor beta receptor binding | MF | 10 | 4 | .0018 | 1 |
| regulation of presynapse organization | BP | 20 | 6 | .0020 | 1 |
| regulation of presynapse assembly | BP | 20 | 6 | .0020 | 1 |

**Table S7b.** Top 10 associated GO-terms for the WGCNA-module “purple” (ventral striatum).

| Term | Ont | N | DE | P.DE | FDR |
|---|---|---|---|---|---|
| regulation of T cell proliferation | BP | 86 | 9 | 4.32E-06 | .0715 |
| regulation of leukocyte cell-cell adhesion | BP | 187 | 13 | 6.83E-06 | .0715 |
| regulation of T cell activation | BP | 197 | 13 | 1.26E-05 | .0881 |
| leukocyte cell-cell adhesion | BP | 205 | 13 | 1.78E-05 | .0932 |
| T cell proliferation | BP | 103 | 9 | 2.38E-05 | .0998 |
| regulation of cell-cell adhesion | BP | 263 | 15 | 2.92E-05 | .1019 |
| regulation of lymphocyte proliferation | BP | 119 | 9 | 7.03E-05 | .1725 |
| negative regulation of T cell proliferation | BP | 30 | 5 | 7.33E-05 | .1725 |
| regulation of mononuclear cell proliferation | BP | 120 | 9 | 7.47E-05 | .1725 |
| regulation of leukocyte proliferation | BP | 123 | 9 | 8.97E-05 | .1879 |

**Table S8a.** GWAS ATLAS results for *IREB2* [retrieved 12.01.2021].

| atlas ID | PMID | Year | Domain | Trait | P-value | N |
|---|---|---|---|---|---|---|
| 4315 | 30643251 | 2019 | Psychiatric | Cigarettes per day | 3.86E-82 | 263954 |
| 3353 | 31427789 | 2019 | Psychiatric | Number of cigarettes previously smoked daily | 6.32E-54 | 90143 |
| 3335 | 31427789 | 2019 | Psychiatric | Light smokers, at least 100 smokes in lifetime | 6.36E-21 | 105610 |
| 20 | 20418890 | 2010 | Psychiatric | Number of cigarettes smoked per day | 4.04E-17 | 38181 |
| 3630 | 31427789 | 2019 | Environment | Illnesses of father: Lung cancer | 2.80E-14 | 355137 |
| 3632 | 31427789 | 2019 | Environment | Illnesses of father: Chronic bronchitis/emphysema | 1.93E-13 | 355137 |
| 4278 | 30804560 | 2019 | Respiratory | FEV1 | 3.78E-13 | 400102 |
| 3640 | 31427789 | 2019 | Environment | Illnesses of mother: Lung cancer | 2.98E-12 | 367939 |
| 3278 | 31427789 | 2019 | Mortality | Father still alive | 1.08E-11 | 375841 |
| 4282 | 30804560 | 2019 | Respiratory | FEV1 | 1.68E-11 | 321047 |
| 3355 | 31427789 | 2019 | Psychiatric | Ever stopped smoking for 6+ months | 5.73E-11 | 93871 |
| 3279 | 31427789 | 2019 | Mortality | Father's age at death | 9.02E-11 | 283990 |
| 3643 | 31427789 | 2019 | Environment | Illnesses of mother: Chronic bronchitis/emphysema | 9.62E-11 | 367939 |
| 4040 | 29906448 | 2018 | Psychiatric | Schizophrenia/Bipolar disorder | 8.75E-10 | 107620 |
| 3277 | 31427789 | 2019 | Environment | Maternal smoking around birth | 2.70E-09 | 331862 |
| 1227 | 28748955 | 2017 | Mortality | Life span | 5.50E-09 | 116279 |
| 4279 | 30804560 | 2019 | Respiratory | FVC | 1.99E-08 | 400102 |
| 4316 | 30643251 | 2019 | Psychiatric | Smoking cessation | 2.14E-08 | 312821 |
| 3982 | 29483656 | 2018 | Psychiatric | Schizophrenia | 3.85E-08 | 105318 |
| 4280 | 30804560 | 2019 | Respiratory | FEV1/FVC ratio | 4.00E-08 | 400102 |
| 11 | 25056061 | 2014 | Psychiatric | Schizophrenia | 1.05E-07 | 82315 |
| 13 | 24280982 | 2014 | Psychiatric | Schizophrenia vs Bipolar disorder | 1.05E-07 | 16381 |
| 3549 | 31427789 | 2019 | Body Structures | Mouth/teeth dental problems: Dentures | 1.27E-07 | 385026 |
| 3887 | 27863252 | 2016 | Immunological | Mean corpuscular hemoglobin (three-way meta) | 4.60E-07 | 172332 |
| 3194 | 31427789 | 2019 | Environment | Number of vehicles in household | 5.73E-07 | 383893 |
| 4283 | 30804560 | 2019 | Respiratory | FVC | 6.77E-07 | 321047 |
| 4284 | 30804560 | 2019 | Respiratory | FEV1/FVC ratio | 8.06E-07 | 321047 |

**Table S8b.** GWAS ATLAS results for *SLC30A8* [retrieved 12.01.2021].

| atlas ID | PMID | Year | Domain | Trait | P-value | N |
|---|---|---|---|---|---|---|
| 4102 | 29403010 | 2018 | Metabolic | Blood sugar | 1.73E-08 | 93146 |
| 4045 | 30054458 | 2018 | Endocrine | Type 2 Diabetes | 1.40E-07 | 659256 |
| 4086 | 30297969 | 2018 | Endocrine | Type 2 Diabetes (adjusted for BMI) | 1.67E-07 | 898130 |
| 4085 | 30297969 | 2018 | Endocrine | Type 2 Diabetes | 5.48E-07 | 898130 |
| 4176 | 30718926 | 2019 | Endocrine | Type 2 Diabetes | 6.10E-07 | 191764 |
| 3469 | 31427789 | 2019 | Metabolic | Impedance measures - Trunk fat mass | 9.85E-07 | 379578 |

**Table S8c.** GWAS ATLAS results for *DDAH2* [retrieved 20.01.2021].

| atlas ID | PMID | Year | Domain | Trait | P-value | N |
|---|---|---|---|---|---|---|
| 1204 | 24390342 | 2014 | Connective Tissue | Rheumatoid Arthritis | 2.0378E-69 | 103638 |
| 1203 | 24390342 | 2014 | Connective Tissue | Rheumatoid Arthritis | 3.6883E-63 | 58284 |
| 2037 | 27723758 | 2016 | Immunological | Immunoglobulin A deficiency | 6.6948E-47 | 6487 |
| 2034 | 27992413 | 2017 | Gastrointestinal | Primary sclerosing cholangitis | 3.6534E-42 | 14890 |
| 4226 | 31015401 | 2019 | Environmental | Thyroid preparations | 1.7121E-31 | 305582 |
| 3904 | 27863252 | 2016 | Immunological | White blood cell count (three-way meta) | 9.8206E-26 | 172435 |
| 3820 | 21980299 | 2011 | Endocrine | Type 1 Diabetes | 1E-25 | 26890 |
| 3187 | 31427789 | 2019 | Skeletal | Standing height | 1.9189E-25 | 385748 |
| 3602 | 31427789 | 2019 | Endocrine | Non-cancer illness code, self-reported: hypothyroidism/myxoedema | 1.1414E-19 | 289307 |
| 3470 | 31427789 | 2019 | Metabolic | Impedance measures - Trunk fat-free mass | 1.8719E-18 | 379507 |
| 3471 | 31427789 | 2019 | Metabolic | Impedance measures - Trunk predicted mass | 2.9617E-18 | 379469 |
| 3412 | 31427789 | 2019 | Skeletal | Sitting height | 5.972E-18 | 385393 |
| 3868 | 27863252 | 2016 | Immunological | White blood cell count (two-way meta) | 2.2674E-17 | 131969 |
| 3892 | 27863252 | 2016 | Immunological | Myeloid white cell count (three-way meta) | 6.5552E-17 | 169219 |
| 3444 | 31427789 | 2019 | Metabolic | Impedance measures - Whole body water mass | 1.8341E-16 | 379835 |
| 3443 | 31427789 | 2019 | Metabolic | Impedance measures - Whole body fat-free mass | 2.5875E-16 | 379804 |
| 4217 | 31015401 | 2019 | Environmental | Drugs used in diabetes | 6.1046E-16 | 305913 |
| 3884 | 27863252 | 2016 | Immunological | Lymphocyte count (three-way meta) | 9.033E-16 | 171643 |
| 3893 | 27863252 | 2016 | Immunological | Sum neutrophil eosinophil count (three-way meta) | 2.3227E-15 | 170384 |
| 3877 | 27863252 | 2016 | Immunological | Granulocyte count (three-way meta) | 2.3317E-15 | 169822 |

| | | | | | | |
|---|---|---|---|---|---|---|
| 3328 | 31427789 | 2019 | Endocrine | Diabetes (diagnosed by doctor) | 1.1639E-14 | 385420 |
| 3872 | 27863252 | 2016 | Immunological | Sum basophil neutrophil count (three-way meta) | 1.279E-14 | 170143 |
| 3895 | 27863252 | 2016 | Immunological | Neutrophil count (three-way meta) | 1.3421E-14 | 170702 |
| 3466 | 31427789 | 2019 | Metabolic | Impedance measures - Arm fat-free mass (left) | 2.5438E-14 | 379653 |
| 3446 | 31427789 | 2019 | Metabolic | Impedance measures - Basal metabolic rate | 2.7498E-14 | 379821 |
| 3889 | 27863252 | 2016 | Immunological | Monocyte count (three-way meta) | 2.8299E-14 | 170721 |
| 3467 | 31427789 | 2019 | Metabolic | Impedance measures - Arm predicted mass (left) | 3.1302E-14 | 379638 |
| 3455 | 31427789 | 2019 | Metabolic | Impedance measures - Leg predicted mass (right) | 3.7464E-14 | 379793 |
| 3454 | 31427789 | 2019 | Metabolic | Impedance measures - Leg fat-free mass (right) | 3.8806E-14 | 379793 |
| 3271 | 31427789 | 2019 | Skeletal | Comparative height size at age 10 | 4.5248E-14 | 380167 |
| 3844 | 27863252 | 2016 | Immunological | Hemoglobin concentration (two-way meta) | 5.4061E-14 | 132596 |
| 4282 | 30804560 | 2019 | Respiratory | FEV1 | 5.6617E-14 | 321047 |
| 208 | 20453842 | 2010 | Connective Tissue | Rheumatoid Arthritis | 8.4057E-14 | 25708 |
| 3891 | 27863252 | 2016 | Immunological | Mean platelet volume (three-way meta) | 8.6069E-14 | 164454 |
| 3463 | 31427789 | 2019 | Metabolic | Impedance measures - Arm predicted mass (right) | 9.4047E-14 | 379716 |
| 3462 | 31427789 | 2019 | Metabolic | Impedance measures - Arm fat-free mass (right) | 9.7426E-14 | 379723 |
| 3856 | 27863252 | 2016 | Immunological | Myeloid white cell count (two-way meta) | 1.0031E-13 | 130268 |
| 3625 | 31427789 | 2019 | Activities | Treatment/medication code: levothyroxine sodium | 1.1682E-13 | 280443 |
| 3529 | 31427789 | 2019 | Nutritional | Never eat eggs, dairy, wheat, sugar: Wheat products | 1.7071E-13 | 384986 |
| 3459 | 31427789 | 2019 | Metabolic | Impedance measures - Leg predicted mass (left) | 1.8863E-13 | 379761 |
| 3458 | 31427789 | 2019 | Metabolic | Impedance measures - Leg fat-free mass (left) | 2.053E-13 | 379766 |
| 3857 | 27863252 | 2016 | Immunological | Sum neutrophil eosinophil count (two-way meta) | 2.9508E-13 | 131409 |
| 3901 | 27863252 | 2016 | Immunological | Red cell distribution width (three-way meta) | 3.0828E-13 | 171529 |
| 3841 | 27863252 | 2016 | Immunological | Granulocyte count (two-way meta) | 3.4015E-13 | 130875 |
| 4278 | 30804560 | 2019 | Respiratory | FEV1 | 3.5176E-13 | 400102 |
| 4080 | 30239722 | 2018 | Metabolic | Waist-hip ratio (adjusted for BMI) | 3.6327E-13 | 694649 |
| 3982 | 29483656 | 2018 | Psychiatric | Schizophrenia | 3.937E-13 | 105318 |
| 4284 | 30804560 | 2019 | Respiratory | FEV1/FVC ratio | 4.4336E-13 | 321047 |
| 4280 | 30804560 | 2019 | Respiratory | FEV1/FVC ratio | 4.6084E-13 | 400102 |
| 3880 | 27863252 | 2016 | Immunological | Hemoglobin concentration (three-way meta) | 4.6242E-13 | 172925 |
| 3859 | 27863252 | 2016 | Immunological | Neutrophil count (two-way meta) | 1.0003E-12 | 131564 |
| 3865 | 27863252 | 2016 | Immunological | Red cell distribution width (two-way meta) | 1.0188E-12 | 131520 |

| | | | | | | |
|---|---|---|---|---|---|---|
| 3836 | 27863252 | 2016 | Immunological | Sum basophil neutrophil count (two-way meta) | 1.0408E-12 | 131031 |
| 3855 | 27863252 | 2016 | Immunological | Mean platelet volume (two-way meta) | 1.1953E-12 | 127230 |
| 3601 | 31427789 | 2019 | Endocrine | Non-cancer illness code, self-reported: diabetes | 1.8686E-12 | 289307 |
| 3848 | 27863252 | 2016 | Immunological | Lymphocyte count (two-way meta) | 1.8817E-12 | 132452 |
| 3899 | 27863252 | 2016 | Immunological | Platelet count (three-way meta) | 2.3443E-12 | 166066 |
| 4085 | 30297969 | 2018 | Endocrine | Type 2 Diabetes | 5.8181E-12 | 898130 |
| 1202 | 24390342 | 2014 | Connective Tissue | Rheumatoid Arthritis | 6.6806E-12 | 22515 |
| 3853 | 27863252 | 2016 | Immunological | Monocyte count (two-way meta) | 1.0206E-11 | 131544 |
| 3552 | 31427789 | 2019 | Respiratory | Blood clot, DVT, bronchitis, emphysema, asthma, rhinitis, eczema, allergy diagnosed by doctor: Asthma | 1.275E-11 | 385822 |
| 3902 | 27863252 | 2016 | Immunological | Reticulocyte count (three-way meta) | 1.4501E-11 | 170641 |
| 3873 | 27863252 | 2016 | Immunological | Sum eosinophil basophil count (three-way meta) | 1.4745E-11 | 171771 |
| 3874 | 27863252 | 2016 | Immunological | Eosinophil count (three-way meta) | 1.865E-11 | 172275 |
| 11 | 25056061 | 2014 | Psychiatric | Schizophrenia | 2.1733E-11 | 82315 |
| 13 | 24280982 | 2014 | Psychiatric | Schizophrenia vs Bipolar disorder | 2.1733E-11 | 16381 |
| 4074 | 30239722 | 2018 | Metabolic | Body Mass Index | 2.8072E-11 | 806834 |
| 3308 | 31427789 | 2019 | Mortality | Long-standing illness, disability or infirmity | 3.1055E-11 | 377498 |
| 2052 | 26691988 | 2016 | Ophthalmological | Age-related Macular Degeneration | 3.278E-11 | 33976 |
| 3447 | 31427789 | 2019 | Metabolic | Impedance measures - Impedance of whole body | 3.3106E-11 | 379792 |
| 3652 | 31427789 | 2019 | Environment | Illnesses of siblings: Diabetes | 3.7124E-11 | 309116 |
| 3436 | 31427789 | 2019 | Metabolic | Weight | 6.2727E-11 | 385473 |
| 3843 | 27863252 | 2016 | Immunological | Hematocrit (two-way meta) | 6.5681E-11 | 132699 |
| 3451 | 31427789 | 2019 | Metabolic | Impedance measures - Impedance of arm (left) | 1.2019E-10 | 379803 |
| 4283 | 30804560 | 2019 | Respiratory | FVC | 1.2738E-10 | 321047 |
| 142 | 25282103 | 2014 | Skeletal | Height | 1.9539E-10 | 253288 |
| 3448 | 31427789 | 2019 | Metabolic | Impedance measures - Impedance of leg (right) | 2.2348E-10 | 379813 |
| 3863 | 27863252 | 2016 | Immunological | Platelet count (two-way meta) | 2.7249E-10 | 127127 |
| 3996 | 29500382 | 2018 | Psychiatric | Nervous feelings (NERV-FEEL) | 2.9739E-10 | 264858 |
| 3879 | 27863252 | 2016 | Immunological | Hematocrit (three-way meta) | 3.1188E-10 | 173039 |
| 3685 | 31427789 | 2019 | Endocrine | Diagnoses - secondary ICD10: E03 Other hypothyroidism | 4.1684E-10 | 244890 |
| 3440 | 31427789 | 2019 | Metabolic | Impedance measures - Weight | 4.1771E-10 | 379840 |
| 3599 | 31427789 | 2019 | Respiratory | Non-cancer illness code, self-reported: asthma | 4.2544E-10 | 289307 |
| 4279 | 30804560 | 2019 | Respiratory | FVC | 4.528E-10 | 400102 |

| | | | | | | |
|---|---|---|---|---|---|---|
| 4082 | 30239722 | 2018 | Metabolic | Waist-hip ratio (adjusted for BMI, female) | 5.0857E-10 | 379501 |
| 3553 | 31427789 | 2019 | Respiratory | Blood clot, DVT, bronchitis, emphysema, asthma, rhinitis, eczema, allergy diagnosed by doctor: Hayfever, allergic rhinitis or eczema | 6.1214E-10 | 385822 |
| 4077 | 30239722 | 2018 | Metabolic | Waist-hip ratio | 7.1194E-10 | 697734 |
| 4086 | 30297969 | 2018 | Endocrine | Type 2 Diabetes (adjusted for BMI) | 7.4293E-10 | 898130 |
| 3838 | 27863252 | 2016 | Immunological | Eosinophil count (two-way meta) | 8.5226E-10 | 131999 |
| 4205 | 31152163 | 2019 | Metabolic | Estimated glomerular filtration rate | 8.5748E-10 | 765348 |
| 3450 | 31427789 | 2019 | Metabolic | Impedance measures - Impedance of arm (right) | 9.4419E-10 | 379786 |
| 3191 | 31427789 | 2019 | Activities | Number of treatments/medications taken | 1.1741E-09 | 386581 |
| 3866 | 27863252 | 2016 | Immunological | Reticulocyte count (two-way meta) | 1.2228E-09 | 130388 |
| 4206 | 31152163 | 2019 | Metabolic | Estimated glomerular filtration rate | 1.2283E-09 | 567460 |
| 3449 | 31427789 | 2019 | Metabolic | Impedance measures - Impedance of leg (left) | 1.2883E-09 | 379807 |
| 4227 | 31015401 | 2019 | Environmental | Immunosuppressants | 1.5537E-09 | 272602 |
| 62 | 26414677 | 2015 | Reproduction | Age at menopause | 2.34E-09 | 69360 |
| 3850 | 27863252 | 2016 | Immunological | Mean corpuscular hemoglobin concentration (two-way meta) | 2.7238E-09 | 132586 |
| 3837 | 27863252 | 2016 | Immunological | Sum eosinophil basophil count (two-way meta) | 2.9274E-09 | 131557 |
| 3903 | 27863252 | 2016 | Immunological | Reticulocyte fraction of red cells (three-way meta) | 3.2798E-09 | 170690 |
| 2733 | 28240269 | 2017 | Cell | CFB - Complement factor B | 4.1835E-09 | 1000 |
| 3551 | 31427789 | 2019 | Cardiovascular | Vascular/heart problems diagnosed by doctor: High blood pressure | 5.244E-09 | 385699 |
| 10 | 23974872 | 2013 | Psychiatric | Schizophrenia | 5.8209E-09 | 32143 |
| 3798 | 29942085 | 2018 | Psychiatric | Worry subcluster | 6.07E-09 | 348219 |
| 4232 | 31015401 | 2019 | Environmental | Anilides | 7.8958E-09 | 179810 |
| 4038 | 29906448 | 2018 | Psychiatric | Schizophrenia | 8.0632E-09 | 87491 |
| 3886 | 27863252 | 2016 | Immunological | Mean corpuscular hemoglobin concentration (three-way meta) | 8.1551E-09 | 172851 |
| 4076 | 30239722 | 2018 | Metabolic | Body Mass Index (female) | 1.1128E-08 | 434794 |
| 2920 | 28240269 | 2017 | Cell | Human-virus - gp41 C34 peptide, HIV | 1.2096E-08 | 1000 |
| 3380 | 31427789 | 2019 | Cardiovascular | Systolic Blood Pressure (automated reading) | 1.2217E-08 | 361402 |
| 4276 | 30833571 | 2019 | Connective Tissue | Carpal tunnel syndrome | 1.4359E-08 | 401656 |
| 4285 | 30804560 | 2019 | Respiratory | PEF | 2.0182E-08 | 321047 |
| 3340 | 31427789 | 2019 | Reproduction | Had menopause (female) | 2.0711E-08 | 175519 |
| 3738 | 31427789 | 2019 | Psychiatric | Depression - Lifetime number of depressed periods | 2.2238E-08 | 57986 |
| 3332 | 31427789 | 2019 | Activities | Taking other prescription medications | 2.5009E-08 | 385236 |
| 3695 | 31427789 | 2019 | Respiratory | Diagnoses - secondary ICD10: J45 Asthma | 2.621E-08 | 244890 |

| | | | | | | |
|---|---|---|---|---|---|---|
| 3186 | 31427789 | 2019 | Metabolic | Hip circumference | 3.1019E-08 | 385887 |
| 4225 | 31015401 | 2019 | Environmental | HMG CoA reductase inhibitors | 3.9029E-08 | 290385 |
| 4081 | 30239722 | 2018 | Metabolic | Waist-hip ratio (adjusted for BMI, male) | 4.0748E-08 | 315284 |
| 2030 | 28067908 | 2017 | Gastrointestinal | Ulcerative colitis | 4.4006E-08 | 45975 |

**Table S8d.** GWAS ATLAS results for *TMEM232* [retrieved 12.01.2021].

| atlas ID | PMID | Year | Domain | Trait | P-value | N |
|---|---|---|---|---|---|---|
| 3553 | 31427789 | 2019 | Respiratory | Blood clot, DVT, bronchitis, emphysema, asthma, rhinitis, eczema, allergy diagnosed by doctor: Hayfever, allergic rhinitis or eczema | 8.61E-13 | 385822 |
| 3552 | 31427789 | 2019 | Respiratory | Blood clot, DVT, bronchitis, emphysema, asthma, rhinitis, eczema, allergy diagnosed by doctor: Asthma | 7.39E-09 | 385822 |
| 3599 | 31427789 | 2019 | Respiratory | Non-cancer illness code, self-reported: asthma | 6.67E-08 | 289307 |
| 4266 | 30929738 | 2019 | Respiratory | Asthma (child-onset) | 1.54E-07 | 314633 |

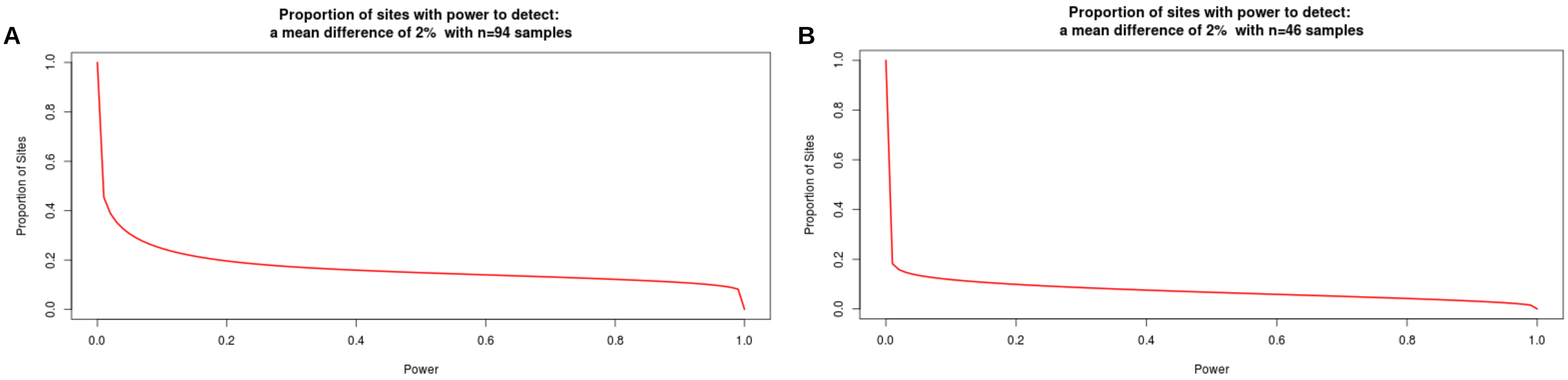


**Figure S1**. Power curve showing the proportion of CpG-sites on the EPIC array with sufficient power to detect a mean methylation difference of 2% for a p-value threshold of $1x10^{-7}$ and a sample size of A) 94 and B) 46.

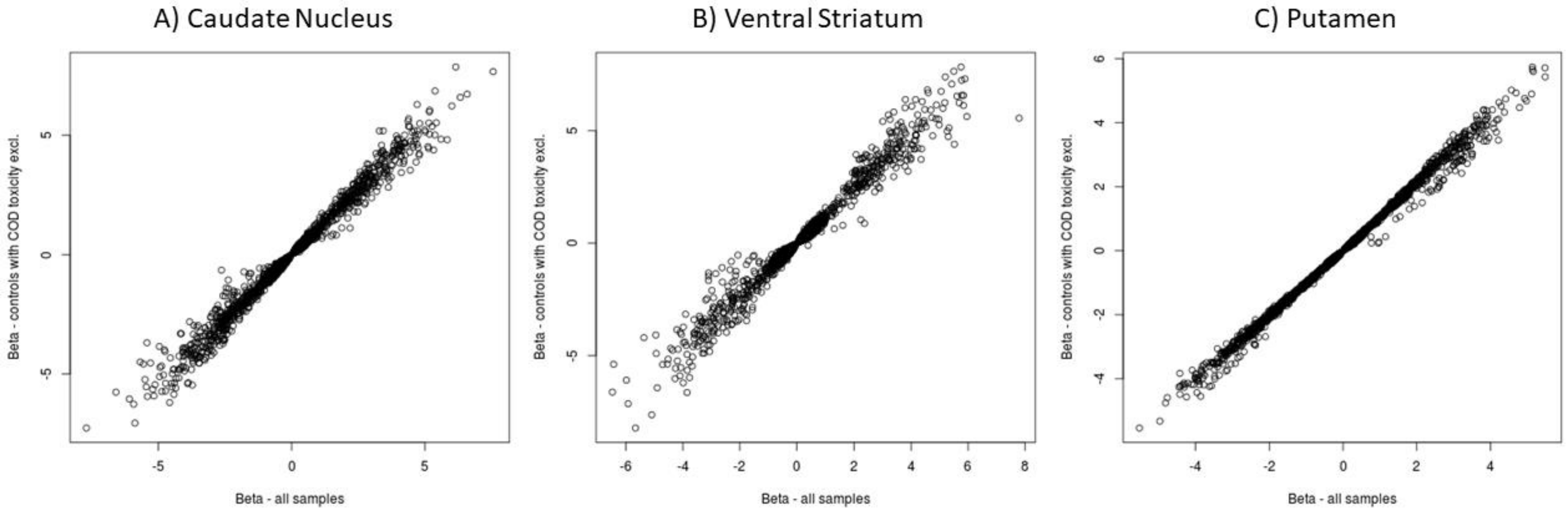


**Figure S2**. Scatterplots of effects for nominally associated CpG-sites from the EWAS in all samples, and from the sensitivity analysis for which control subjects with cause of death toxicity were excluded.

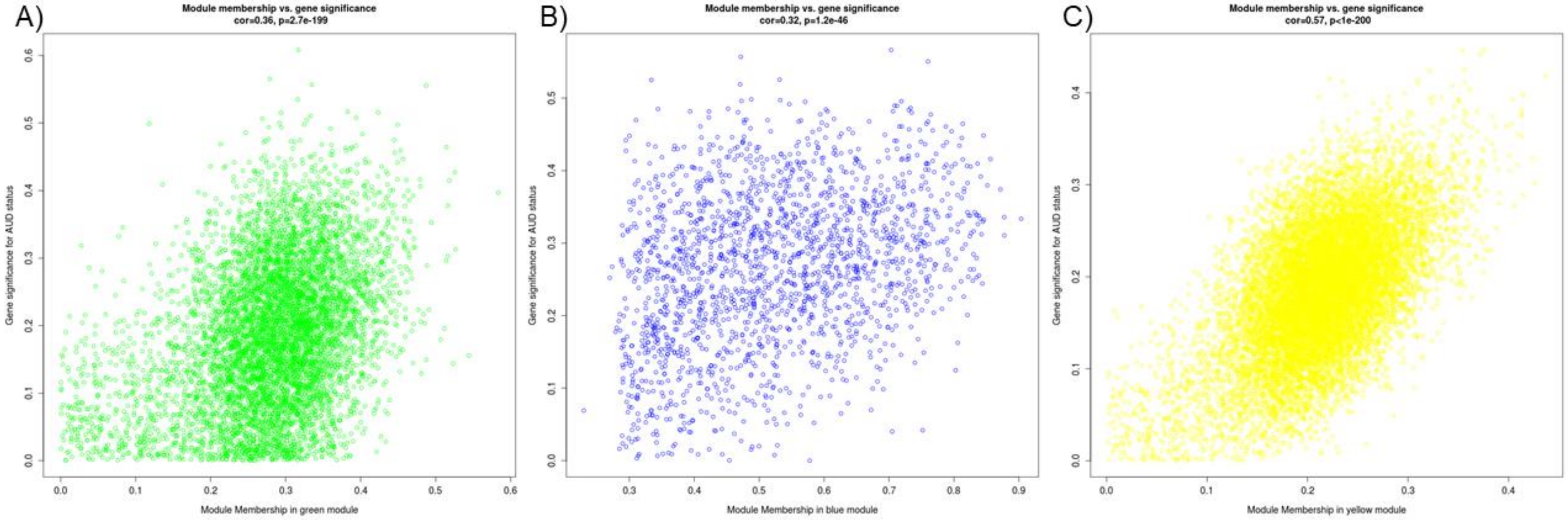


**Figure S3**. Scatterplots of CpG-site significance and module membership, of WGCNA modules most strongly associated with AUD in A) anterior cingulate cortex, B) Brodmann Area 9, and C) Putamen.
